# Estrogen upregulates NF-κB and TNF signaling in B cells to cause sex and age differences in antibody responses to seasonal influenza vaccination

**DOI:** 10.1101/2025.05.22.25328179

**Authors:** Han-Sol Park, Anna Yin, Weiqiang Zhou, Eliane F.E. Wenstedt, Anne Jedlicka, Amanda Dziedzic, Annie Werner, Katherine Z.J. Fenstermacher, Huifen Li, Patrick J. Shea, John S. Lee, Patricia Gearhart, Robert W. Maul, Sjoerd A.A. Van Den Berg, Richard E. Rothman, Hongkai Ji, Sean X. Leng, Andrew Pekosz, Sabra L. Klein

## Abstract

Sex differences in the humoral immune responses to the seasonal quadrivalent influenza vaccine (QIV) in young adults (YA; 18-49yo) or high dose QIV in old adults (OA; 75+yo) were analyzed to evaluate how age-related changes in steroids impact sex differences in B cell responses.

Among YAs, females had greater H3N2, but not H1N1, neutralizing antibody titers and greater proportions of hemagglutinin (HA)+CD19+ B cells and HA+ memory B cells than males through 28 days post-vaccination (DPV), which was not observed among OAs. Machine learning algorithms illustrated that baseline (0 DPV) steroids, including 17-hydroxyprogesterone, estrogens, and testosterone, as well as HA+CD19+ B cells and HA+ antibody-secreting B cells (ASCs) were major predictors of subsequent seroconversion at 28 DPV, particularly in YA. Single cell RNA sequencing demonstrated CD19+ B cells from YA females had greater transcriptional activity at 7 DPV than YA males, with upregulation of genes with estrogen-response elements (EREs) along NF-κB-mediated TNF signaling pathways in B cell subsets, including naïve, memory, and ASCs, which was mitigated in OA. Estradiol treatment of ASCs from YA females, but not males, increased the number and size of HA+IgG+ ASCs and expression of genes with EREs along the NF-κB-mediated TNF signaling pathway, which were inhibited by an estrogen receptor antagonist. Pharmacological inhibition of either NF-κB or TNF signaling blocked the ability of E2 to upregulate antibody secretion from YA female ASCs. This study provides mechanistic insights into estrogen mediated increases in influenza vaccine-induced antibody responses among reproductive-aged females as compared with age-matched males and suggests a role for estrogen signaling in the reduction of sex differences in vaccine-induced immunity with old age.

**One Sentence Summary:** Estrogenic activity in B cells causes greater influenza vaccine-induced immunity in young adult females than age-matched males, with these sex differences being mitigated in old adults.

## INTRODUCTION

Across the life course, biological sex and age are fundamental modifiers of immunity to infectious diseases and the response to vaccination. Females tend to mount stronger immune responses than males (*1–3*), but these sex differences in immunity are not constant and change with age. There is an intersection between sex and age, whereby the impact of aging on the immune system differs in males and females (*4, 5*). The implications of the interaction between sex and age are clearly demonstrated by the epidemiology and clinical manifestations of respiratory viral diseases, such as influenza and COVID-19 (*6, 7*). Influenza A viruses (IAVs) have caused four of the five pandemics during the past century, with seasonal influenza epidemics resulting in approximately 650,000 deaths worldwide, with people 65+ years of age being at a higher risk of severe outcomes (*8*). The 2019-2020 influenza season was one of the last major influenza seasons, with a notable decline in influenza activity during the COVID-19 pandemic, followed by a resurgence starting in 2022-2023.

Of the few human influenza vaccine trials that have considered sex differences in vaccine outcomes, in adults of reproductive ages (18-49 years), females have greater antibody titers (i.e., either neutralizing or hemagglutination inhibition [HAI] titers) than males following receipt of the seasonal influenza vaccine (*9–12*). The sex difference in antibody responses is influenced by both age and pre-existing immunity to selected IAV and influenza B virus (IBV) strains (*11, 13*). During the 2008-2009 influenza season, adult females, for example, produced more neutralizing antibodies against the H3N2 and IBV vaccine virus strains, but not against the H1N1 vaccine virus strain (*11*). During the 2017-2018 influenza season, adult females mounted more robust neutralizing responses to H3N2 vaccine virus compared to males (*14*). A recent meta-analysis of 19 randomized controlled trials of influenza vaccination revealed a slightly higher rate of seroconversion in females compared to males, with the sex differential effect being greatest among individuals <65 years of age (*15*).

Among individual 65+ years of age, antibody titers to both the standard and high dose seasonal influenza vaccine are also greater in females than males (*15, 16*), with greater season to season durability of immunity against influenza among older females than males, particularly in response to H3N2 and IBV (*13*). Both younger age and greater concentrations of circulating estradiol (E2) are associated with greater monovalent 2009 H1N1 vaccine-induced immunity (*9*), suggesting that declining gonadal steroids, and not just chronological aging, could reduce immunity following vaccination. Estrogen therapy in postmenopausal women increases antibody responses to seasonal influenza vaccines (*17*). Limited data also suggest that sex differences in influenza-vaccine induced antibody responses involve the effects of testosterone on lipid biosynthesis in males (*11*). Missing from the current literature is a deep interrogation into how biological sex, age, and gonadal steroids impact the molecular and cellular pathways in B cells to cause sex differences in antibody responses following vaccination.

Herein, we conducted parallel studies of young adult (YA) healthcare workers ages 18-49 at the Johns Hopkins Hospital System receiving the standard dose of the quadrivalent influenza vaccine (QIV) and community-dwelling older adults at least 75 years of age (OA) in Baltimore, Maryland receiving the recommended high dose QIV. We show that estrogen impacts signaling through NF-κB and TNF pathways in B cells, resulting in increased vaccine-induced antibody responses in YA females compared to males. In OA, these sex differences are mitigated, in part because of reduced estrogen signaling in B cells from OA females.

## RESULTS

### Patient demographics

During the 2019-2020 influenza season, 82 young adult (YA) healthcare workers (HCW) at the Johns Hopkins Health System (JHHS) between the ages of 18 and 49 years old were enrolled prior to receipt of the required seasonal standard dose inactivated quadrivalent influenza vaccine (QIV) and completed sample collection (**Table 1**). The YA cohort comprised of 43 males and 39 females, with an overall average age of 30 years. The YA participants consisted of 50% White, 17% Black or African American, 17% Asian, 2% American Indian or Alaskan Native, 10% other, and 4% with missing race data. Among the YA females, 26 (68%) were on birth control (BC), none were pregnant, none had a hysterectomy, and none were on hormone replacement therapy (HRT).

**Table 1.** Participant demographics and clinical comorbidities for the young adult and old adult cohorts during the 2019-2020 influenza vaccination season.

|  | Young adult cohort<br>18-49 years old |  |  |  | Old adult cohort<br>75+ years old |  |  |  |
| --- | --- | --- | --- | --- | --- | --- | --- | --- |
|  | Male | Female | Total | P-value* | Male | Female | Total | p-value |
| **n, %** | 43 (52) | 39 (47) | 82 (100) | - | 37 (44) | 47 (56) | 84 (100) | - |
| **Age, mean (SD)** | 29.3 (6.2) | 30.9 (7.5) | 30.0 (6.9) | 0.29 | 85.5 (5.3) | 83.7 (5.1) | 84.5 (5.3) | 0.13 |
| **BMI, mean (SD)** | 25.8 (3.8) | 26.6 (5.4) | 26.2 (4.6) | 0.42 | 27 (4) | 25.5 (5.1) | 26.2 (4.6) | 0.08 |
| **Race, n (%)** |  |  |  | **0.02** |  |  |  | 1.0 |
| White | 19 (44) | 22 (56) | 41 (50) |  | 32 (86) | 38 (81) | 70 (83) |  |
| Black/African American | 4 (9) | 10 (26) | 14 (17) |  | 2 (5) | 3 (6) | 5 (6) |  |
| Asian | 12 (28) | 2 (5) | 14 (17) |  | 0 (0) | 0 (0) | 0 (0) |  |
| American Indian or Alaska Native | 2 (5) | 0 (0) | 2 (2) |  | 0 (0) | 0 (0) | 0 (0) |  |
| Other | 4 (9) | 4 (8) | 8 (10) |  | 0 (0) | 0 (0) | 0 (0) |  |
| N/A | 2 (5) | 1 (3) | 3 (4) |  | 3 (8) | 6 (13) | 9 (11) |  |
| **Hispanic/Latino, n (%)** | 8 (19) | 7 (18) | 15 (18) | 1.0 | - | - | - | - |
| **Breastfeeding, n (%)** | - | 1 (3) | 1 (1) | - | - | - | - | - |
| **Hysterectomy, n (%)** | - | 0 (0) | 0 (0) | - | - | 7 (15) | 7 (8) | - |
| **Immunosuppressive medication, n (%)** | 2 (5) | 1 (3) | 3 (4) | 1.0 | 0 (0) | 0 (0) | 0 (0) | - |
| **Autoimmune disease, n (%)** | 2 (5) | 3 (8) | 5 (6) | 0.66 | 20 (54) | 36 (77) | 56 (67) | **0.03** |
| **Cardiovascular disease, n (%)** | 1 (2) | 3 (8) | 4 (5) | 0.34 | 8 (22) | 7 (15) | 15 (18) | 0.42 |
| **Asthma, n (%)** | 2 (5) | 3 (8) | 5 (6) | 1.0 | 0 (0) | 3 (6) | 3 (4) | 0.25 |
| **Reproductive disease, n (%)** | 0 (0) | 4 (10) | 4 (5) | - | - | - | - | - |
| **Cancer, n (%)** | 0 (0) | 1 (3) | 1 (1) | 0.47 | 16 (43.2) | 17 (36.2) | 33 (39.3) | 0.65 |
| **H3N2 seroprotection at 0 DPV, n (%)** | 11 (28) | 22 (63) | 33 (45) | **0.003** | 14 (44) | 16 (37) | 30 (40) | 0.57 |
| **H3N2 seroprotection at 28 DPV, n (%)** | 34 (87) | 33 (94) | 67 (91) | 0.44 | 22 (69) | 32 (74) | 54 (72) | 0.59 |
| **H3N2 seroconversion, n (%)** |  |  |  | 0.27 |  |  |  | 0.66 |
| Non-responder (< 4 FC) | 14 (36) | 16 (46) | 30 (41) |  | 20 (63) | 29 (67) | 49 (65) |  |
| Responder (≥ 4 FC) | 25 (64) | 19 (54) | 44 (59) |  | 12 (37) | 14 (33) | 26 (35) |  |
| **nAb data missing, n (%)** | 4 (9) | 4 (10) | 8 (10) |  | 5 (14) | 4 (8) | 9 (11) |  |

\*comparison between sexes and within an age group

We concurrently enrolled community-dwelling old adults (OAs) in Baltimore above 75 years of age who had not yet received the high-dose seasonal QIV. A total of 84 participants were enrolled and completed sample collection, comprising of 37 males and 47 females with an overall average age of 84.5 years old (**Table 1**). The OA cohort was predominately White (83%, n=70) with 6% (n=5) identifying as Black or African American and 11% (n=9) missing race data. Among the OA females, 8% (n=7) had a previous hysterectomy and none were on HRT.

### Influenza vaccine compositional changes 2015-2020 season

In 2019, the egg-based QIV composition consisted of A/Brisbane/02/2018(H1N1)pdm09-like virus, A/Kansas/14/2017(H3N2)-like virus, B/Colorado/06/2017-like virus (B/Victoria/2/87 lineage), and B/Phuket/3073/2013-like virus (B/Yamagata/16/88 lineage) in accordance with the World Health Organization’s (WHO) vaccine recommendations for the Northern Hemisphere (*18*). From 2015 to 2019, the H1N1 vaccine component changed three times whereas H3N2 vaccine component changed four times (**Fig. 1A**). To determine how these changes in the vaccine components affected antigenic sites, the hemagglutinin (HA) trimer structures from A/H1N1 (A/California/07/2009 PDB:5WKO) and A/H3N2 (A/Switzerland/9715293/2013 PDB:6PDX) were visualized with Protein Imager and amino acid changes between the vaccine components were mapped. The H1 HA protein used in the 2019 influenza vaccine had 23 amino acid changes relative to the vaccine strain used in 2015, with many of them located in or near antigenic sites (**Fig. 1B**) In contrast, the H3 HA protein had 38 amino acid changes with 12 of those specific sites having changed twice (e.g., amino acids 187, 175 and 160) during this period (**Fig. 1C**). These data indicate that individuals vaccinated with the recommended Northern Hemisphere influenza vaccine formulation were exposed to H3 components that had more antigenic site changes than the H1 component.

**Figure 1:**
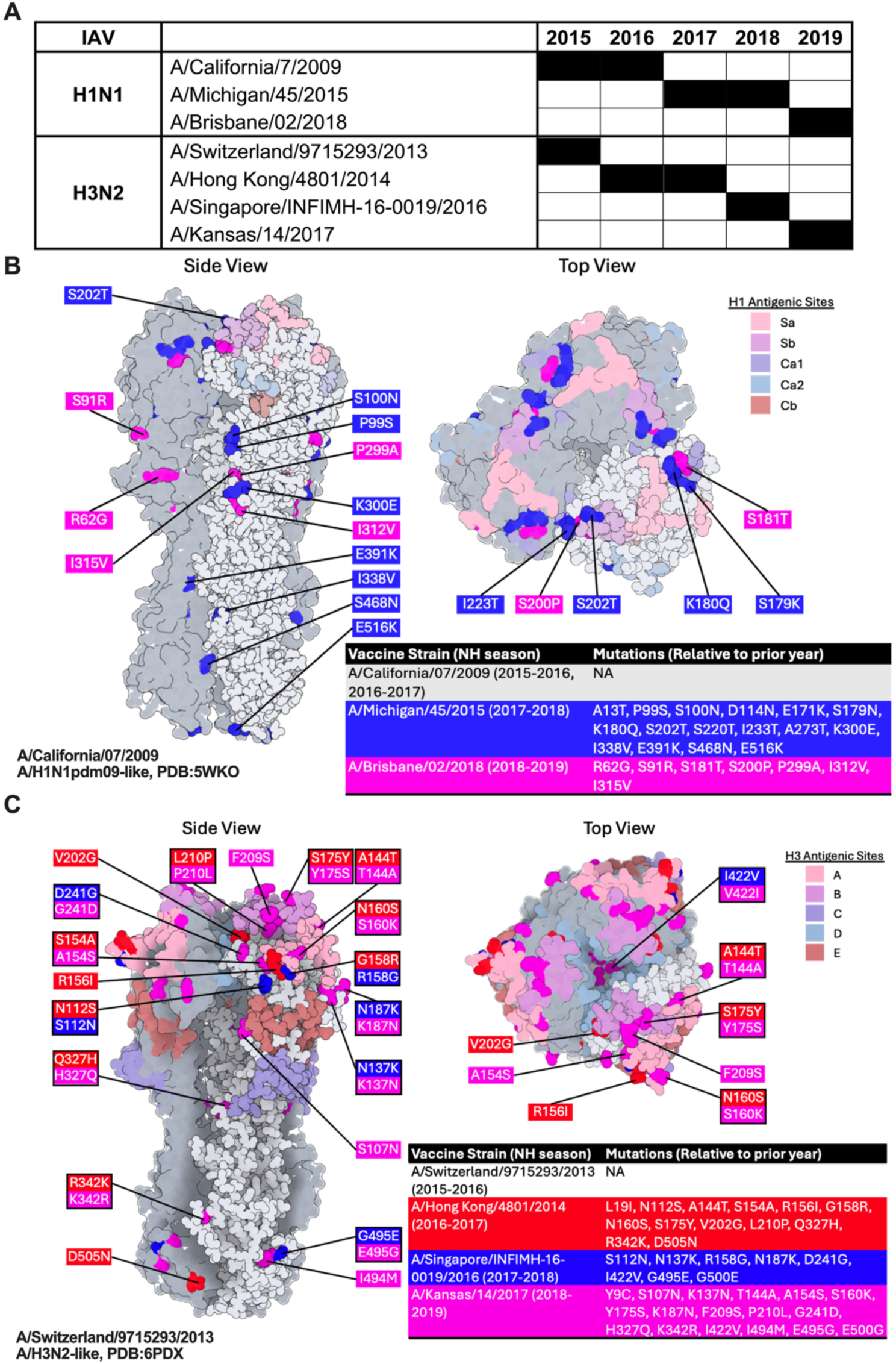
Strain changes in the Northern Hemisphere quadrivalent influenza vaccine (QIV) formulation from 2015-2019. (**A**) H1N1 and H3N2 components of the Northern Hemisphere (NH) inactivated influenza vaccine from 2015-2019. (**B**) HA trimers from A/H1N1 (A/California/07/2009 PDB:5WKO) and (**C**) A/H3N2 (A/Switzerland/9715293/2013 PDB:6PDX) were visualized using Protein Imager. The major antigenic sites are shaded in pastel colors and amino acid changes in the vaccine strains are indicated with the specific amino acid substitution and the vaccine strain it was present in indicated. Boxes with two or more amino acid changes indicate multiple changes during this timeframe at that specific site. The H1N1 components collectively had fewer amino acid changes in antigenic sites compared to the H3N2 components.

### YA females have greater pre- and post-vaccination neutralizing antibody titers against H3N2 than YA males, with sex differences mitigated in OA

Sex differences in neutralizing antibody (nAb) responses to IAVs are typically limited to pandemic strains of H1N1 and seasonal strains of H3N2 (*9, 13, 19*). Regardless of age group or sex, all participants (100%) had nAb titers against H1N1 influenza that were well above that for seroprotection (>1:40 titer, log2) prior to vaccination, suggesting strong pre-existing immunity against H1N1 in 2019-2020 (**Fig. S1A,C**). Despite strong pre-existing nAb titers against H1N1 prior to vaccination, males and females similarly increased nAb titers against H1N1 between 0 days post vaccination (DPV) and 28 DPV with no statistically significant sex differences observed, regardless of age. The average fold change in H1N1 nAb titers between 28 and 0 DPV was 1.7 for YA females and 3.1 for YA males (p=0.13). Among OA, the average fold change in H1N1 nAb titers was 2.6 for females and 2.1 for males (p=0.17). These data suggest that although influenza vaccination was able to induce nAb titers against H1N1 for YA and OA, most participants (n=61, 82% of YA and n=51, 68% of OA) did not seroconvert for H1N1 (< 4-fold change). Even when YA and OA were separated between non-responders (< 4-fold change) and responders (>4-fold change), sex differences in nAb titers against H1N1 were not observed (**Fig. 2A-B**).

**Figure 2:**
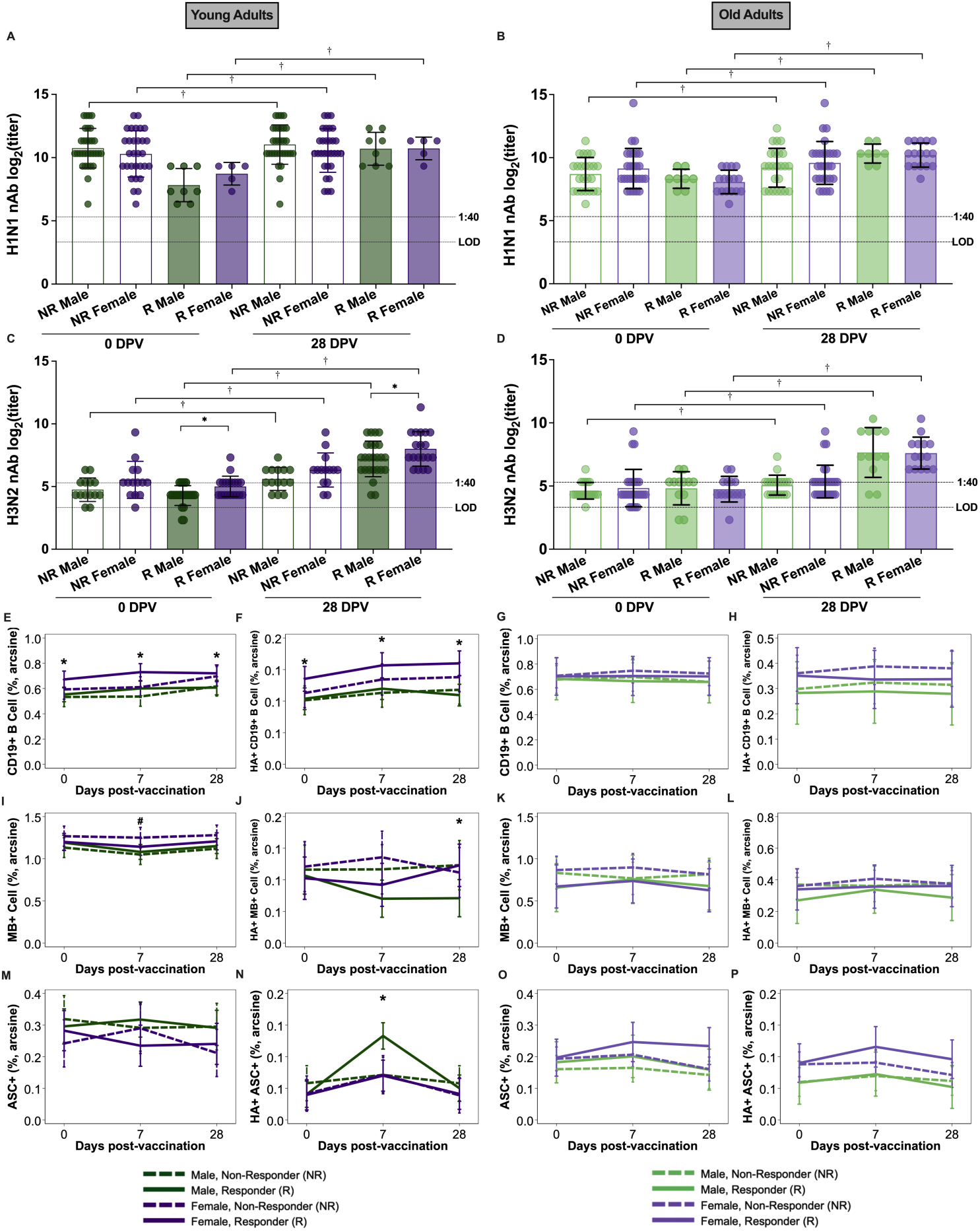
Among young, but not old, adult vaccine responders, females had greater influenza-specific immune responses than males. Vaccine responders (R) were defined as individuals that showed a >4-fold rise in nAb titers 28 days post vaccination (DPV) relative to their pre-vaccination titer (0 DPV). Non-responders (NR) were individuals with <4-fold rise in nAb titers after vaccination. In response to H1N1 (**A-B**), most young (79% of male and 85.7% of female participants) and old (75% of male and 62.8% female participants) adult participants were NR because their pre-vaccination nAb titers were sufficiently high and did not have at least a 4-fold change in nAb titer following vaccination. In response to H3N2, 36% of male and 45.7% of female young adult participants and 62.5% of male and 67.4% of female old adult participants were NR, suggesting less pre-existing immunity to H3N2 than H1N1 vaccine viruses (**C-D**). Dashed lines represent the limit of detection (LOD) and log_2_ (1:40 titer) for seroprotection. Proportions of total and HA+ CD19+ B cells (**E-H**), memory B cells (MB; **I-L**), and antibody-secreting cells (ASC; **M-P**) were quantified by flow cytometry in young and old adult participants. Data collected across multiple timepoints were analyzed by linear mixed effects regression models with an interaction term for sex, timepoint, H3N2 seroconversion status, adjusting for continuous age. Young and old adults were analyzed independently unless specified. * and # represent p<0.05 for sex differences among R or NR, respectively and † represents p<0.05 for nAb increases over time.

There were significantly lower pre-existing nAb titers against H3N2 than H1N1 influenza at 0 DPV. YA females had greater H3N2 nAb titers both prior to and 28 DPV than YA males, with no sex differences in H3N2 nAb titers observed among OA (**Fig. S1B,D**). When YA and OA were separated into non-responders and responders, sex differences in nAb titers were only apparent among YA responders, in which females had greater nAb titers against H3N2 than males (**Fig. 2C-D**). The average fold change in H3N2 nAb titers between 28 DPV and 0 DPV was 6.6 for males and 8.9 for females among YA (p=0.55). In OA, the average fold change for H3N2 nAb titers was 4.8 for males and 3.9 for females (p=0.59). In response to H3N2, 36% of male (n=14) and 45.7% of female (n=16) YA participants and 62.5% of male (n=20) and 67.4% of female (n=29) OA participants did not seroconvert for H3N2 (< 4-fold change). Together, these data suggest that YA female responders were driving the sex differences in nAb responses against H3N2. To determine if birth control (BC) use could impact nAb titers to either H1N1 or H3N2, we compared YA females who reported using BC with those not using BC. BC use did not impact titers of nAb against either H1N1 (**Fig. S1E**) or H3N2 (**Fig. S1F**).

### YA females maintain greater frequencies of HA+ B cells than YA males before and after vaccination, with sex differences mitigated in OA

We used flow cytometry to measure total and HA+ B cell frequencies in peripheral blood mononuclear cell (PBMC) samples among YAs and OAs collected before and after seasonal influenza vaccination (**Fig. S2**). Because responders were driving sex differences in nAb responses, we separated responders and non-responders based on H3N2 nAb seroconversion.

Prior to and after vaccination, YA females maintained significantly higher frequencies of total and HA+ CD19+ B cells compared to YA males (**Fig. S3A-B**), which was driven by responders who seroconverted (**Fig. 2E-F**). In contrast, OA females and males had similar frequencies of total and HA+ CD19+ B cells, regardless of seroconversion status (**Fig. S3C-D** and **Fig. 2G-H**). Total memory B cell frequencies (defined as CD3e^-^CD14^−^CD16^−^CD19^+^IgD^-^CD71^-^) were greater among YA females than males at 7 DPV (F**ig. S3E**), which was caused by non-responder females with elevated pre-existing immunity (**Fig. 2I**). In contrast, HA+ memory B cells were greater in PBMCs from YA females than males at 28 DPV and this was specific to responders who seroconverted (**Fig. S3F** and **Fig. 2J**). In contrast, OA females and males had similar frequencies of total and HA+ memory B cells, regardless of seroconversion status (**Fig. S3G-H** and **Fig. 2K-L**). Antibody secreting cells (ASCs or plasmablasts; defined as CD3e^-^ CD14^−^CD16^−^ CD19^+^IgD^-^CD71^+^CD20^-^CD38^+^) are the primary circulating B cell type that produces antibody, with peak frequencies occurring within one week after vaccination (*20*). Changes in frequences of total ASCs were not observed over time in either YA or OA males or females (**Fig. S3I-K** and **Fig. 2M-O**). In contrast, proportions of HA+ ASCs peaked at 7 DPV in YA (**Fig. S3J**) but not OA (**Fig. S3L**), which was driven primarily by YA male responders (**Fig. 2N**). OA males and females, regardless of seroconversion, did not exhibit a peak in total or HA+ ASCs at 7 DPV (**Fig. 2O-P**).

To determine if birth control (BC) use among YA females could impact B cell frequencies, we compared proportions of total and HA+ B cell subpopulations between YA females using BC to those not using BC. Consistent with nAb titers, there were no differences in the frequencies of either total or HA+ CD19+ B cells (**Fig. S4A-B**), memory B cells (**Fig. S4C-D**), or ASCs (**Fig. S4E-F**). These data highlight that regardless of whether YA females have endogenous circulating or receive exogenous steroid hormones, the frequencies of B cells, including HA+ B cells, are similar before and after vaccination.

Age-associated B cells (ABCs), also referred to as atypical memory B cells, accumulate in circulation with increasing age, and are often associated with immunosenescence (*21*).

Previous studies illustrate that older females tend to have greater numbers of ABCs than age matched males or younger aged females and males, particularly in autoimmune disease patients(*21*). We explored frequences of ABCs (defined as CD3e^-^ CD14^−^ CD16^−^ CD19^+^CD21^-^CD11c^+^Tbet^+^) in a subset of YA and all OA vaccinees prior to vaccination and observed that OA had significantly greater frequencies of total and HA+ ABCs than YA (**Fig. S5A-B**). To determine if ABC frequencies changed following vaccination in OA, we measured ABCs in OA males and females before and after vaccination and observed no changes in frequencies between the sexes (**Fig. S5C-D**).

### Concentrations of steroids, precursors, and metabolites differ based on sex and age

We have shown that gonadal steroids correlate with sex and age differences in influenza vaccine induced immunity in humans and mice (*9*). In mice, estradiol (E2) causes greater influenza vaccine induced immunity and protection in both young and old aged females by increasing germinal center B cell immune responses (*22*). Different from our previous studies (*9, 22*), steroids in this study were measured by mass spectrometry in plasma samples collected prior to vaccination, which allowed for more accurate assessment of steroid biosynthesis (**Fig. 3A**).

**Figure 3:**
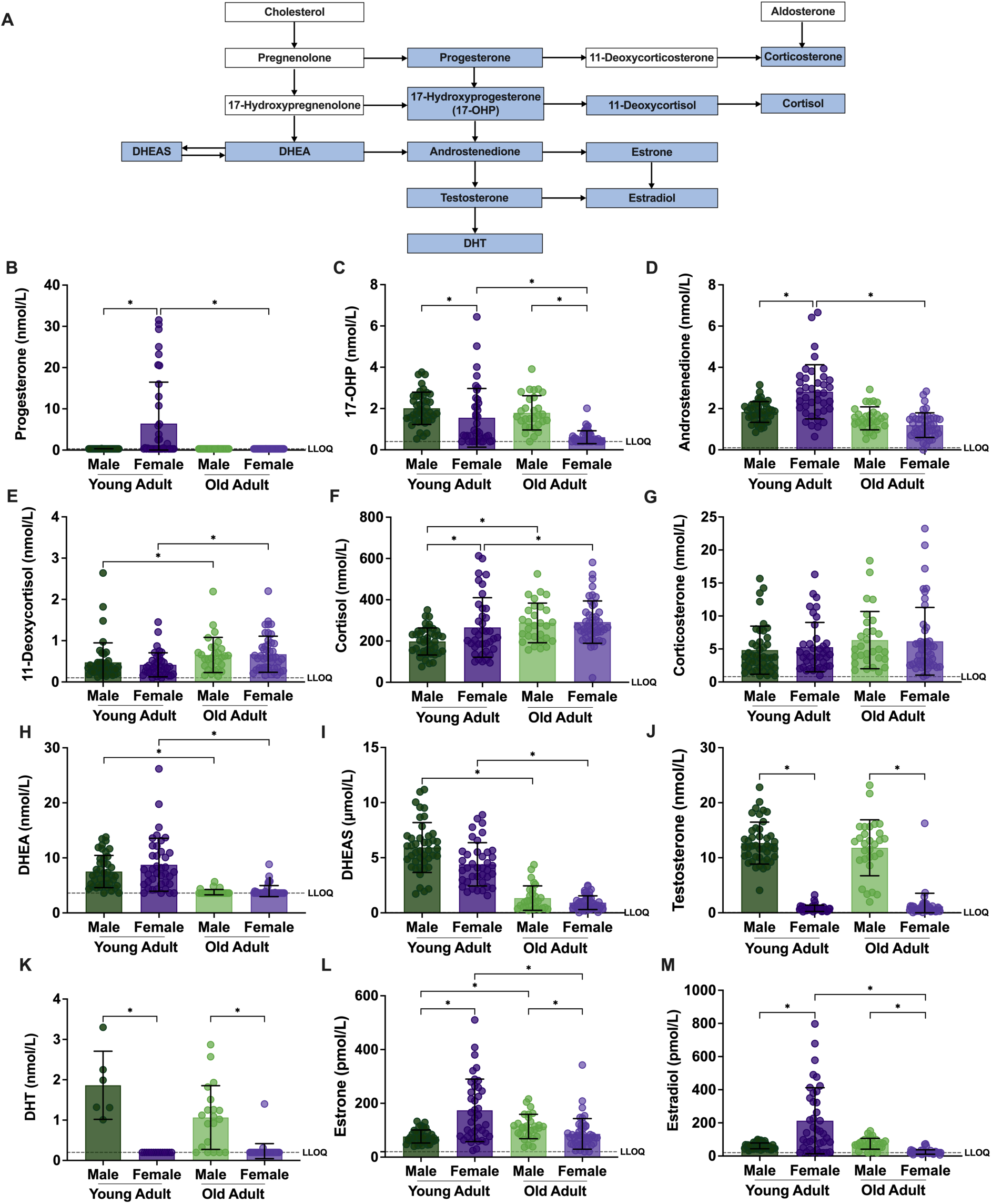
Prior to vaccination, young and old adults had distinct steroid profiles among male and female participants. (**A**) The steroid hormone biosynthesis pathway, with those measured by mass spectrometry highlighted in blue. **(B-M)** Baseline concentrations (mmol/L, nmol/L, or pmol/L) of steroids as well as their precursors and metabolites, were measured from peripheral blood samples collected prior to vaccination (0 DPV) and compared by sex and age using Kruskal-Wallis with Benjamini-Hochberg post-hoc corrections. Dashed lines represent the lower limit of quantification (LLOQ). * represents sex or age differences at padj<0.05.

Progesterone is a precursor for most steroids, and YA females had greater concentrations than either YA males or OAs of either sex (**Fig 3B**). 17-hydroxyprogesterone (17-OHP; **Fig. 3C**) and androstenedione (**Fig. 3D**) concentrations also were greater in YA females than males, with age-related reductions apparent among females, but not males. Both 11-deoxycortisol (**Fig. 3E**) and cortisol (**Fig. 3F**) increased with age in both males and females. Corticosterone was measured and was not different among YA and OA males and females (**Fig. 3G**).

Dehydroepiandrosterone (DHEA) (**Fig. 3H**) and dehydroepiandrosterone sulfate (DHEAS; **Fig. 3I**) are steroid precursors that showed an age-associated decline in both males and females. Androgens, including testosterone (**Fig. 3J**) and dihydrotestosterone (DHT; **Fig. 3K**), were found in greater concentrations among males than females. In contrast to our measurements using immunoassays that are more prone to cross-reactivity (*9*), an age-associated decline in testosterone and DHT concentrations among males was not observed, which has been reported in other studies using mass spectrometry (*23, 24*).

Concentrations of estrogens, including estrone (**Fig. 3L**) and E2 (**Fig. 3M**), were greater in YA females than males, with an age-associated decline occurring specifically in females. In contrast, an age-related increase in estrogens was observed in males, similar to other studies utilizing mass spectrometry (*25*). This is further demonstrated by the stronger positive correlations observed between estrone and estradiol concentrations in YA females (ρ=0.88, p<0.001) as compared to YA males (ρ=0.60, p<0.001); whereas among OA, males had a stronger correlation (ρ=0.72, p<0.001) than females (ρ=0.65, p<0.001).

### Baseline sex steroid concentrations and HA+ CD19+ B cell responses predict seroconversion in YA more than OA

To determine if H3N2 seroconversion status could be predicted using baseline (0 DPV) measures from vaccinees, we applied a machine learning algorithm, known as random forest (RF) modeling (*26*), to steroid hormone, nAb, total and HA+ CD19+ B cell subset data collected prior to vaccination for YA and OA. Complete baseline datasets without missing data were available for 27 non-responders and 44 responders among YA while OA had 44 non-responders and 19 responders. To address potential class imbalance issues, we downsampled the datasets to the minor class, resulting in 27 responders and non-responders for YA and 19 non-responders and responders for OA (**Fig. S6A-B**). Using a 5-fold cross-validated random forest modeling approach to predict a binary seroconversion status (responder or non-responder), which was repeated across 10 different seeds, we found that the gonadal steroid precursor, 17-OHP, % of HA+ CD19+, % of HA+ ASC, estrone, and testosterone were among the top 5 features most important for classifying seroconversion status for YA, as represented by the average importance (**Fig. 4A**). Because estrone concentrations in YA were highly correlated with estradiol concentrations prior to vaccination, particularly among YA females, estradiol may be as useful as estrone in classifying seroconversion status among YA. Averaging model performance metrics across 10 seeds, the seroconversion models for YA were able to classify the testing data with an accuracy of 60.63%, area under the receiver operator curve (AUROC) of 0.61, and area under the precision-recall curve (AUPRC) of 0.54 (**Fig. S6A, C**). In OA, the feature importance analysis following RF modeling for predicting seroconversion revealed that % MB+, % ASC+, % CD19+ B cells, and estrone were among the top features needed for classification (**Fig. 4B**). However, the RF models did not have strong model performance, with an average accuracy of 54%, AUROC of 0.48, and AUPRC of 0.46 (**Fig. S6A, D**), suggesting that baseline antibody, cellular, and steroid data are insufficient to predict subsequent seroconversion for OA.

**Figure 4:**
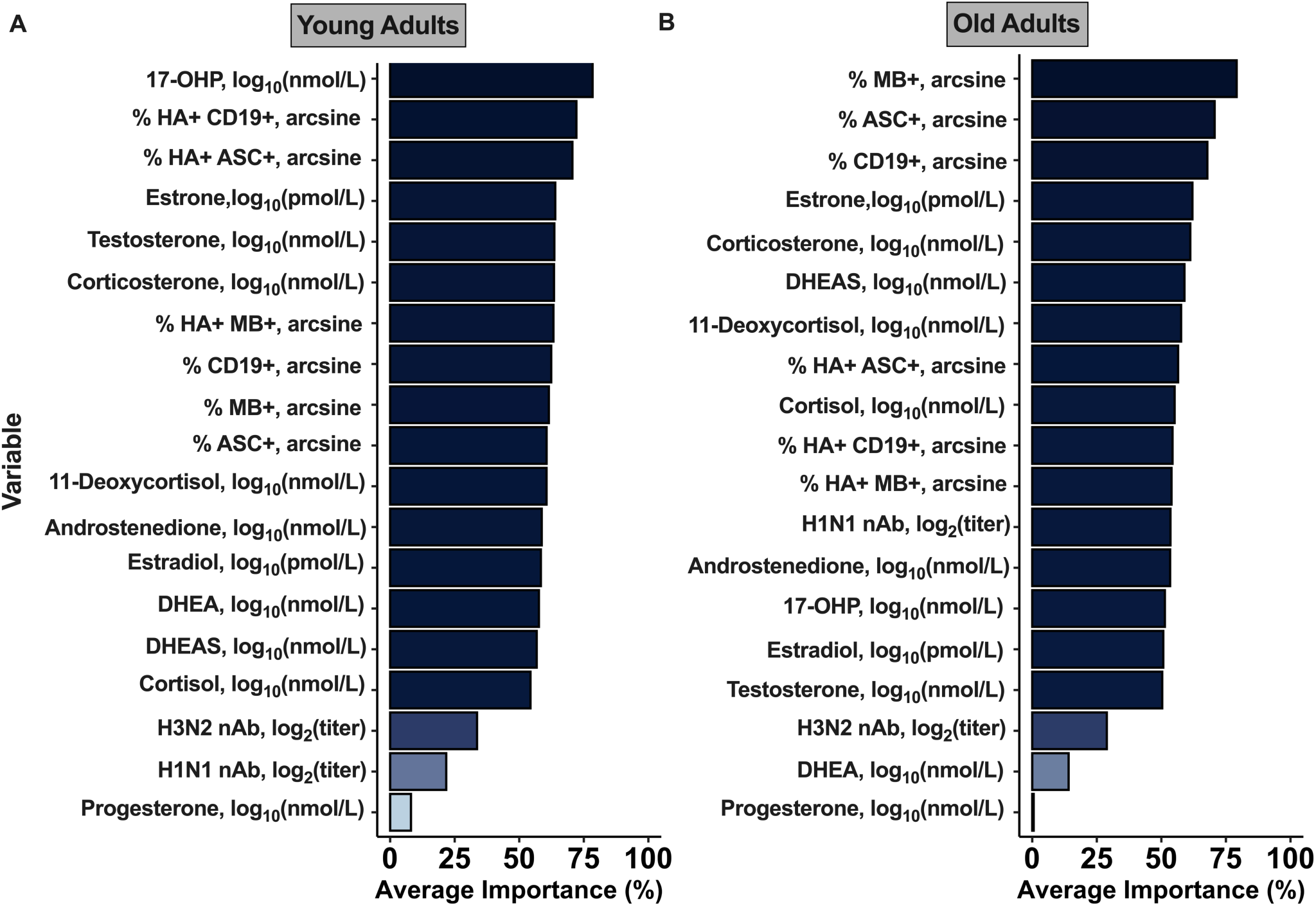
5-fold cross-validated random forest modeling predicted seroconversion using pre-vaccination (0 DPV) measurements among young and old adults. Datasets were downsampled and stratified by outcome variable before being partitioned, with 70% used for training the 5-fold cross-validated random forest models with 5 repeats and averaged across 10 distinct seeds. (**A**) Variable importance plot (VIP) for the random forest model predicting seroconversion among young adults. **(B)** Variable importance plot (VIP) for the random forest model predicting seroconversion among old adults. Average feature importance was calculated by averaging the mean decrease accuracy (%) for 10 distinct seeds. Bars are colored and ranked by descending average feature importance.

To ensure that the RF model predictions were able to be attributed to the steroid, nAb, and cellular measures at baseline and not by random chance, we permuted the seroconversion labels of the downsampled data, performed five-fold cross-validated random forest modeling, and averaged across 10 different seeds. As expected, the RF modeling performance metrics decreased, with the average prediction accuracy of YA dropping to 50.63% and that of OA dropping to 45% (**Fig. S6B, E-F**). AUROC and AUPRC also dropped to 0.48 and 0.47 for YA, respectively, and 0.38 for both metrics for OA (**Fig. S6B, E-F**). These findings suggest that sex steroid concentrations and pre-existing levels of HA+ CD19+ B cell subpopulation frequencies may be predictive of post-vaccination seroconversion status among YA, but not OA, which further underscore the age-associated role of sex steroids, including estrogens, in shaping influenza vaccine immunity.

### Age-associated differences in transcriptional activity of CD19+ B cells prior to vaccination

To explore whether the age and sex differences of nAb responses were related to the underlying transcriptional activity of CD19+ B cells, we identified a subset (**Fig. S7**) of YA and OA males and females with steroid concentrations, antibody responses, and CD19+ B cell frequencies, including HA+ B cell subsets, that were reflective of the larger cohort (**Fig. 2**). For the sample subset, we FACS sorted CD19+ B cells prior to single cell RNA sequencing (scRNAseq).

While scRNAseq analyses comparing YA and OA participants at 7 and 28 DPV were confounded by the different doses of QIV administered as per CDC recommendations, we explored age-related transcriptional activity of B cells at ‘baseline’ prior to vaccination. Principal component analysis of pseudobulk gene expression revealed that prior to vaccination, the gene expression profiles of YA and OA were distinct for both males and females (**Fig. S8A**).

Differential gene expression (DGE) analysis comparing age groups revealed that males had more differentially expressed genes (DEGs) than females (**Fig. S8B**). Gene set enrichment analysis (GSEA) of ranked DEGs revealed that the hallmark pathways most significantly enriched (padj<0.05) in OA were tumor necrosis factor (TNF) signaling via NF-κB and inflammatory response pathways, with *SOCS3, DUSP5, NR4A3, RGSI, CD69,* and *TNSF9* being the leading genes of these pathways, for both males and females (**Fig. S8C-H**). These data point to an ‘inflammaging’ of CD19+ B cells in both males and females, which has been linked to dysregulated B cell metabolism and activity, including antibody production (*27, 28*). Both YA males and females had enrichment of the hallmark interferon alpha (IFNα) signaling pathway (padj<0.05, **Fig. S8C-D**), which included a number of interferon stimulated genes (ISGs; **Fig. S8I-J**) that have not been well characterized in B cells outside of production of autoantibodies (*29*), but may be critical in vaccine-induced antibody production.

### Sex differences in CD19+ B cell transcriptional activity may be regulated by estrogens, particularly in YA

To evaluate sex differences among YA or OA, we conducted PCA of pseudobulk gene expression profiles from FACS-sorted CD19+ B cells collected at 0, 7, or 28 DPV (**Fig. 5**). Sex differences in pseudobulk gene expression were not observed prior to vaccination or at 28 DPV for either YA or OA. Pseudobulk gene expression patterns for males and females were distinct at 7 DPV among YA, but not OA (**Fig. 5A-B**). Focusing on the 7 DPV timepoint to further investigate the sex differences, we found that CD19+ B cells from YA females had enrichment of hallmark TNF signaling via NF-κB and estrogen response signaling pathways (padj<0.05), whereas CD19+ B cells of YA males had enrichment of hallmark pathways related to metabolism and IFNα signaling (padj<0.05, **Fig. 5C**). At 7 DPV, there were no hallmark pathways enriched in OA females, but several of the same pathways related to metabolism and IFNα signaling that were enriched in YA males also were enriched in OA males (padj<0.05, **Fig. 5D**).

**Figure 5:**
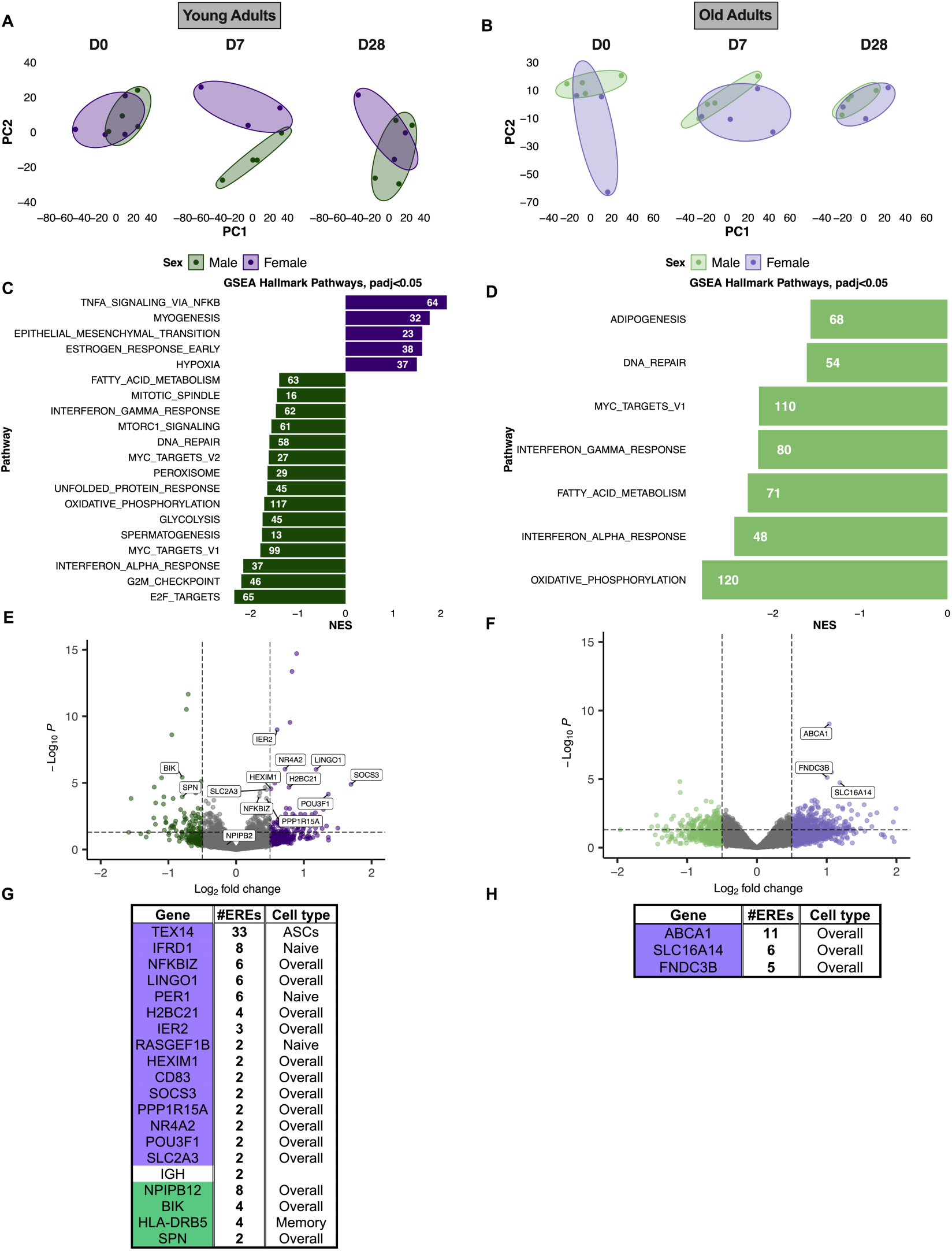
Sex differences in transcriptional activity between 7- and 0-days post vaccination (DPV) were apparent among young, but not old, adults. (**A-B**) Principal component analysis (PCA) plots of pseudobulk gene expression by DPV, colored by sex (purple for females and green for males). (**C-D**) Statistically significant Hallmark gene set enrichment (GSEA) pathways (padj<0.05) are shown along with number of leading-edge genes associated with each pathway and normalized enrichment scores (NES). (**E-F**) Volcano plots depict differentially expressed genes (DEGs) at padj<0.05 and log_2_FC<|0.05|, with selected DEGs for estrogen response element (ERE) analysis labeled. (**G-H**) ERE analyses were conducted using EREFinder using the top DEGs from pseudobulk analyses with ERE motifs (AGGTCAnnnTGACCT) in the promoter region (-5000 base pairs [TSS] to +3000 base pairs [TSS]).

To further investigate the role of estrogens in the regulation of gene expression in CD19+ B cells, we used EREFinder (*30*) to identify and quantify putative estrogen response elements (EREs) in the promoter region (-5000 to +3000 base pairs) of top DEGs between males and females at 7 DPV. Among the DEGs between YA males and females at 7 DPV, we identified several genes containing EREs in their promoter regions, most of which were among the upregulated genes of YA female CD19+ B cells (**Fig. 5E**). On the other hand, three DEGs containing EREs were identified among OA females, with none found in the genes upregulated in the CD19+ B cells of OA males at 7 DPV (**Fig. 5F**). Quantification of DEGs containing EREs by EREFinder further revealed that upregulated genes in CD19+ B cells of YA females contained more EREs than those of YA males (**Fig. 5G**). CD19+ B cells of OA females had fewer DEGs containing EREs than YA females, with OA males having no DEGs that contained EREs (**Fig. 5H**). Together, these data suggest that sex differences of CD19+ B cell activity at 7 DPV may be due to an estrogenic regulation of gene expression via estrogen receptor binding to the consensus ERE motifs, particularly for YA females.

### Age-associated sex differences in signaling pathways in B cell subpopulations

To define B cell subpopulations from scRNAseq of FACS-sorted CD19+ B cells, Seurat Uniform Manifold Approximation and Projection (UMAP) clustering of single cells was used with manual cell type annotations of canonical markers (*31*). With the exception of ABCs, which were only identified in OA, and double negative (DN) and transitional (T1-3) B cells, which were only identified in YA, ASCs, memory B cells (MB), and several populations of naïve B cells were identified in both YA and OA participants, regardless of sex (**Fig. S9A-D**). By proportion, naïve and MB cell populations were the greatest for YA and OA alike. Person to person heterogeneity was observed in the proportions of B cell subsets, but this was not distinguishable based on the sex, age, or time point of sample collection (**Fig. S9E-J**).

Age-related sex differences were most pronounced at 7 DPV from our pseudobulk gene expression analysis; therefore, we focused our single gene set enrichment analyses (ssGSEA) at that timepoint to evaluate which CD19+ B cell subpopulations may be driving hallmark pathway enrichment in YA and OA, particularly the signaling, immune, and metabolic gene sets. Z-scaled enrichment scores (ES) were calculated for CD19+ B cell subpopulations for each sex among YA and OA, respectively (**Fig. 6A-B**). For both YA and OA, we observed that ASCs, naïve, and MB cells were consistently driving the enrichment of these hallmark pathways. Inflammation and estrogen response pathways were more enriched in YA females than YA males, with the reversed pattern observed among OA (i.e., greater enrichment in OA males than female) across diverse B cell subpopulations (**Fig. 6A-B**). Further, the greater enrichment of the hallmark estrogen response early and TNF signaling via NF-kB pathways across several CD19+ B cell subpopulations of YA females as compared to YA males was consistent with our pseudobulk-level findings (**Fig. 5**) and the greater concentrations of estrone and estradiol among YA females (**Fig. 3L-M**). Differential pathway analysis across CD19+ B cell subpopulations further revealed that the hallmark TNF signaling via NF-κB pathway was consistently among the top differentially enriched pathways between males and females across most subpopulations, with greater normalized enrichment scores (NES) in females than males, regardless of age group (padj<0.05; **Fig. 6C-D**). These data continue to highlight the role of estrogen in inflammatory responses in CD19+ B cell gene expression following influenza vaccination, including in ASCs.

**Figure 6:**
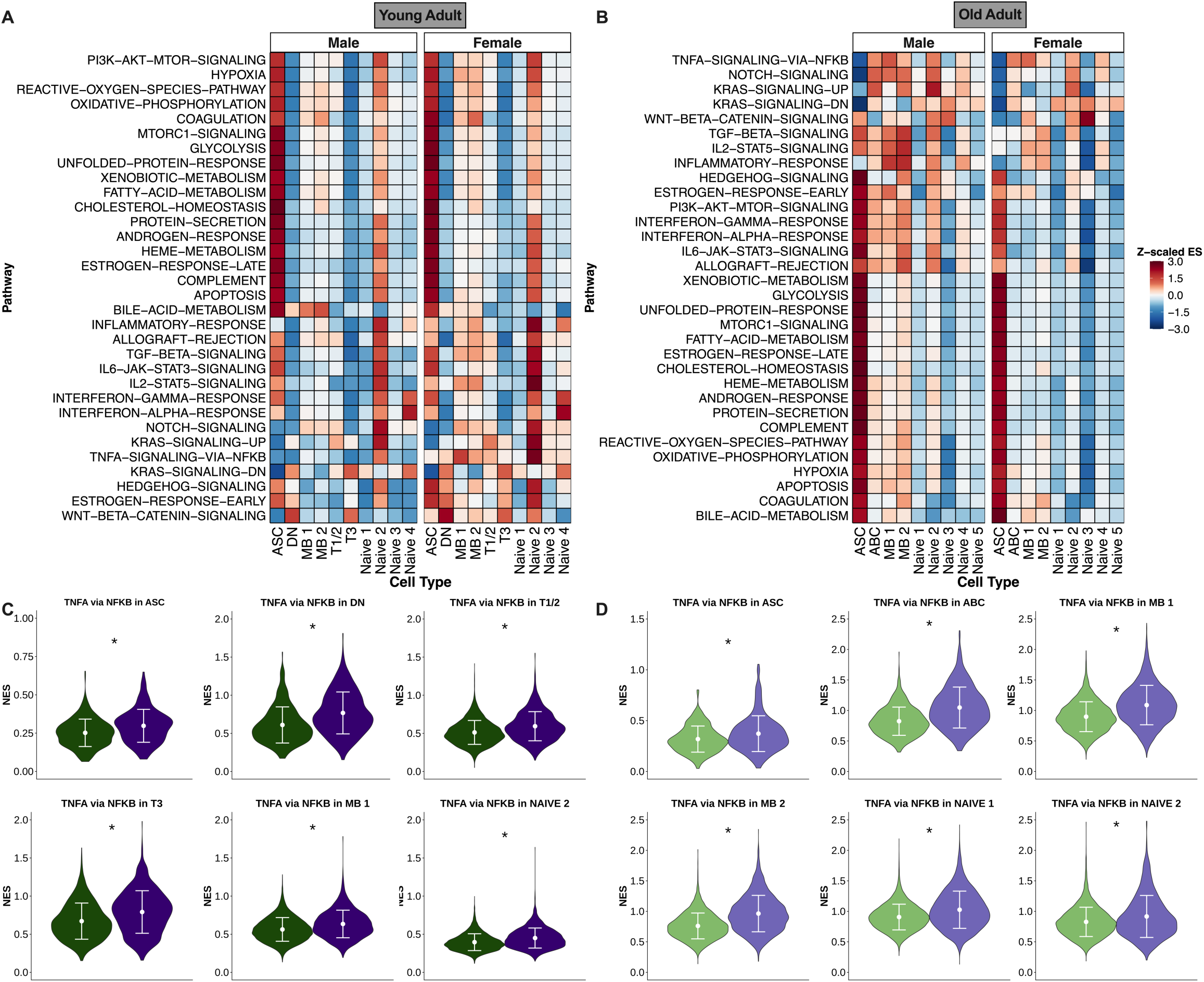
Single sample gene set enrichment analysis (ssGSEA) at 7 DPV in FACS sorted CD19+ B cells from young and old adults. (**A-B**) Z-scaled enrichment scores (ES) were calculated at the single cell level using the ‘escape’ package. Mean Z-scaled ES values are shown in the heatmaps by cell types and sex for young and old adults, respectively. Hallmark gene sets pertaining to signaling, immune, and metabolic pathways are shown. (**C-D**) Cell types with |log2FC|> 0.2 and padj<0.05 for sex differences in normalized enrichment scores (NES), as calculated by differential pathway analysis, with the TNF signaling via NFKB gene set being one of the most sex differential pathways in the greatest number of CD19+ B cell subsets.

### Females have greater HA+ IgG production by ASCs

We next sought to develop an *ex vivo* model to study the mechanisms of sex and age differences in antibody secretion following HA stimulation. We stimulated PBMCs from YA and OA females and males with H1, H3, or pooled H1/H3 antigen and analyzed HA-specific IgG production in samples collected prior to vaccination and 7 and 28 DPV. YA females, and to a lesser extent OA females, had greater numbers HA-specific IgG+ cells than age-matched males at 7 DPV only (**Fig. 7A-D**). Greater numbers of HA-specific IgG+ cells were primarily in response to H3 and to a lesser extent H1 antigen (**Fig. S10A-H**). In addition, the spot sizes corresponding to the amount of HA-specific IgG secreted per cell in YA and OA were greater in females than males (**Fig. 7E-F**), which was primarily caused by greater IgG secretion in response to the H3 than H1 antigen (**Fig. S10I-L**). These data suggest that when cell numbers are normalized across ages and sexes, ASCs from females, regardless of age, have a greater propensity to produce antibody in response to HA than cells from males.

**Figure 7:**
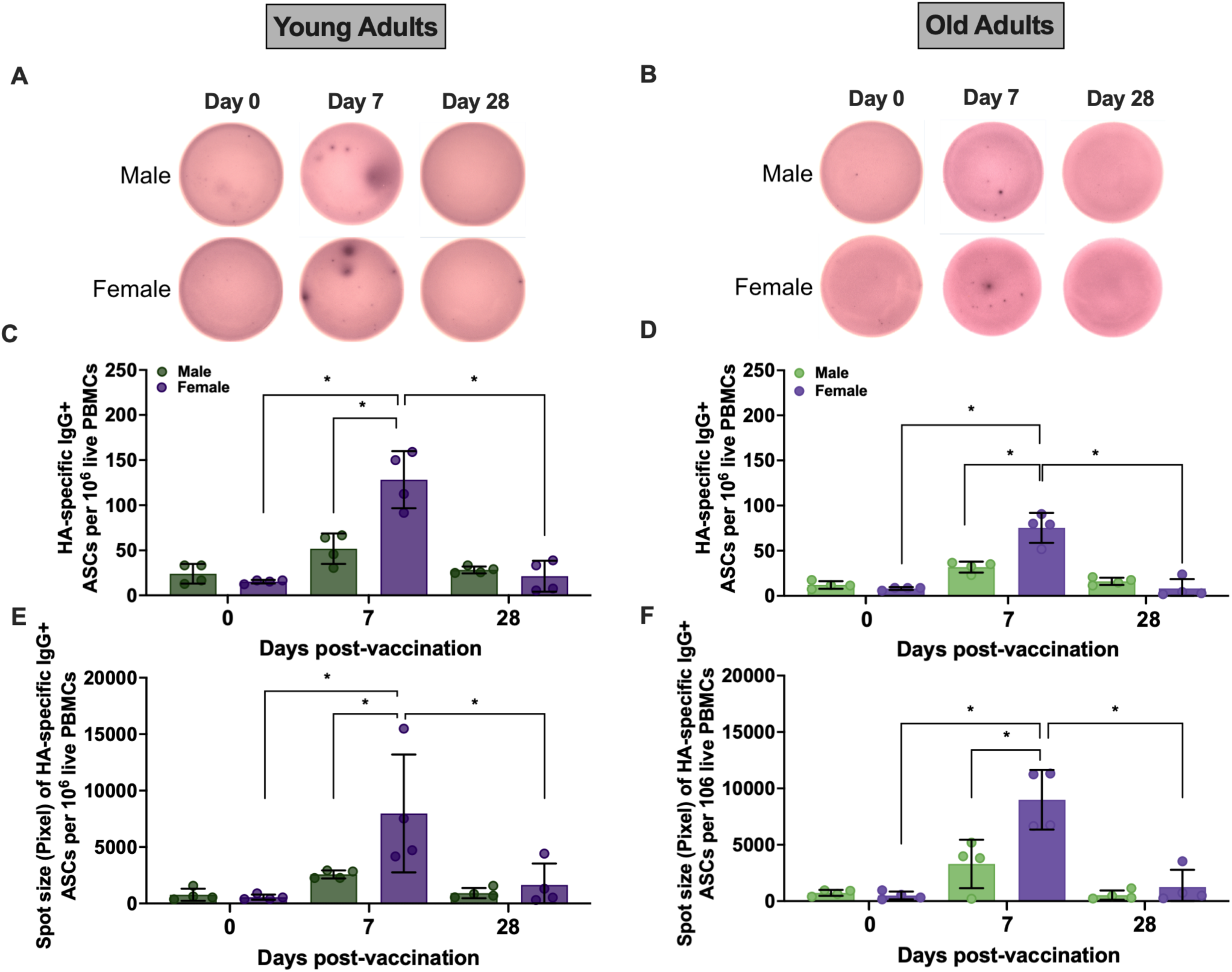
Signaling through estrogen receptors enhanced production of HA-specific IgG by ASCs from females compared to males, independent of age. PBMCs from equal numbers of male and female participants were analyzed for HA-specific IgG production at multiple time points. (**A, B**) Representative images of antibody-secreting cells (ASCs) producing HA-specific IgG from PBMCs collected male and female participants prior to and 7- and 28-days post-vaccination (DPV). Spots represent HA-specific IgG+ cells. Quantification of the number of HA-specific IgG+ cells in PBMCs from young adults (**C**) and old adults (**D**). (**E, F**) Quantification of spot sizes corresponding to HA-specific IgG+ in young and old adults. A total of 1 × 10⁶ PBMCs from 4 male and 4 female participants were used. Data were analyzed by one-way ANOVA with Tukey’s post-hoc corrections. * represents p < 0.05.

### Estradiol enhances HA+ IgG secretion via TNF and NF-κB signaling

Studies in mice highlight that elevated E2 concentrations cause greater influenza vaccine-induced antibody responses in adult females than males and account for the age-related decline in immunity among females (*9, 22*). With age-related differential estrogen concentrations and signaling in B cell subpopulations, including ASCs, we sought to explore if E2 signaling through the ER in B cells was a cause of higher antibody titers in YA females than males, which was mitigated in OA. PBMCs collected at 7 DPV were stimulated with HA antigens and treated overnight with either media alone (untreated), E2, or tamoxifen (TAM; ER antagonist) prior to E2. In PBMCs from YA females (**Fig. 8A**), and to a lesser extent from OA females (**Fig. 8B**), treatment with E2 significantly increased the number (**Fig. 8C-D**) and size (**Fig. 8E-F**) of HA-specific IgG+ cells over treatment with media alone. There was no effect of E2 on cells from males. Pre-treatment with TAM before E2 significantly reduced both the number (**Fig. 8C-D**) and size (**Fig. 8E-F**) of HA-specific IgG+ cells in females but not males among both YA and OA. These data illustrate that ER activation causes increased antibody secretion by B cells in females, with the cells of OA females maintaining the propensity to respond to estrogen.

**Figure 8:**
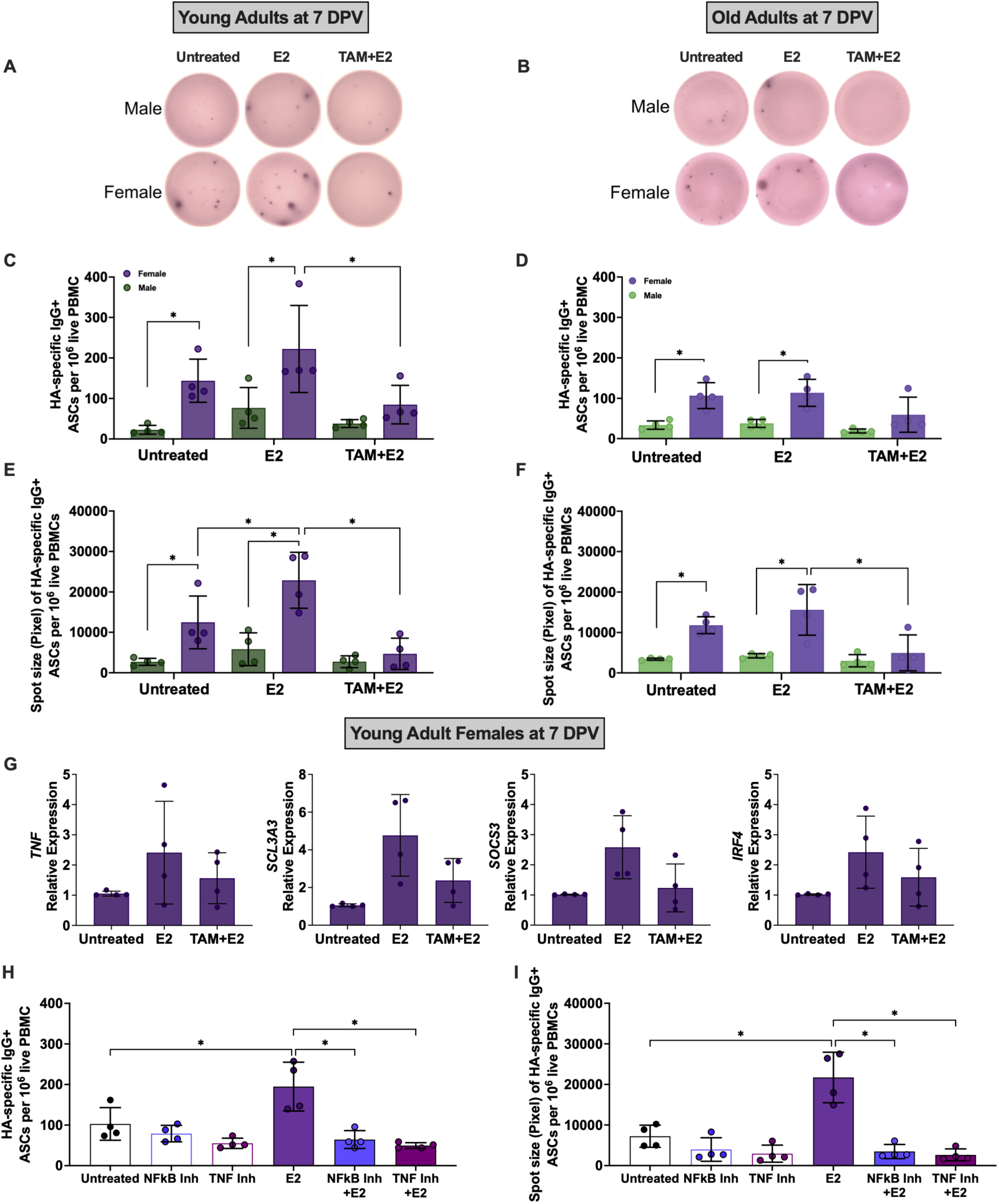
Estradiol enhances HA-specific IgG secretion through the TNFα signaling via NFκB pathway. PBMCs from young adult (YA) and old adult (OA) female and male participants were analyzed for HA-specific IgG production at 7 days-post vaccination (DPV). (**A, B**) Representative images of spots after overnight stimulation of PBMCs with either media alone (untreated), 100nM 17β-estradiol (E2), or 1μM tamoxifen (TAM) treatment prior to E2. (**C, D**) Quantification of the number of HA-specific IgG+ cells following treatments. (**E, F**) Quantification of spot sizes for HA-specific IgG+ cells after treatments. (**G**) qRT-PCR of *TNF*, *SCL3A3*, *SOCS3,* and *IRF4* expression in untreated, E2, and TAM treatment prior to E2 in sorted CD19+ B cells from YA females at 7 DPV. (**H**) Quantification of the number of HA-specific IgG+ cells of following treatment with either media alone (untreated) or 100nM E2 in the presence or absence of either 1μM NFκB inhibitor (Inh; Bay 11-7082) or 1μg/mL TNFα inhibitor (Etanercept). Inhibitors were incubated with PBMCs for 1 hour prior to E2 treatment. A total of 1 × 10⁶ PBMCs 4 female participants were used. Data were analyzed by one-way ANOVA with Tukey’s post-hoc corrections. * represents p < 0.05.

Estrogen and TNF/NF-κB signaling pathways are both upregulated in B cells from female YAs (**Fig. 5**) but whether E2 causes increased B cell activity via TNF/NF-κB signaling is not clear. To determine whether increased secretion of IgG in the presence of E2 in YA females was associated with increased expression of DEGs along the TNF signaling via NF-κB pathway, we sorted CD19+ B cells collected at 7 DPV from YA females and evaluated the expression of genes identified as having putative EREs (**Fig. 5G**), including *TNFA*, *SCL3A3*, *SOCS3*, and *IRF4.* The expression of all these genes was upregulated in the presence of E2 and downregulated by antagonism of ER with TAM (**Fig. 8G**). To determine if E2 caused greater antibody secretion in YA females through either TNF or NF-κB, we pharmacologically inhibited either NF-κB or TNFα in either the presence or absence of E2. E2 could not increase either the number (**Fig. 8H**) or size (**Fig. 8I**) of HA+IgG+ cells from YA females in the absence of either NF-κB or TNF at doses that had no effect of cell viability (**Table S1**). These data illustrate that estrogenic enhancement of antibody secretion requires signaling through NF-κB and TNF in response to vaccine antigen.

## DISCUSSION

Sex differences in the humoral immune responses to vaccines are rarely considered and even more rarely mechanistically explored in humans. Sex differences in antibody responses are not static and can change with aging, suggesting a role for gonadal steroids. We and others have shown that females of reproductive ages have greater influenza vaccine-induced nAb titers than age-matched males following receipt of either seasonal influenza vaccine H3N2 and IBV strains or novel pandemic influenza vaccines (e.g., 2009 H1N1) (*9, 11, 13*). In the current study, YA females had greater H3N2, but not H1N1, nAb titers, and greater proportions of HA+CD19+ B cells and HA+ memory B cells than males, which was not observed among OAs. These sex differences were primarily caused by the vaccine responders, not the non-responders who had high nAb titers prior to vaccination. These data highlight that elevated pre-existing immunity masks sex differences in humoral immunity to IAV antigens, at least among reproductive aged individuals. It has been demonstrated that human H3N2 influenza virus evolves antigenically at a higher rate than either H1N1 or IBV (*32*). This translates into reduced pre-existing immunity to H3N2 in the population because antigenic site mutations in H3N2 occur and spread globally faster, leading to more frequent changes in the H3N2 vaccine component. Among OA, sex differences post vaccination were not observed for any of the immune parameters, regardless of seroconversion status. There are different selective pressures in serum from YA versus OA recently vaccinated against influenza that results in specific amino acid changes at antigenic sites having different effects on escape from vaccine-induced immunity(*33*). Age-dependent antibody specificity differences might also contribute to the lack of sex differences seen in the OA, particularly if they were responding to vaccines that were stimulating strong preexisting B cell responses, which do not show sex differences.

Antibody secreting cells are the early producers of antigen-specific antibody that generally peak within one week following influenza vaccination and decline thereafter (*20*). Frequencies of HA+ ASCs did not exhibit a characteristic peak in the OAs but among YAs, males had greater frequencies than females at 7 DPV. When cell numbers were normalized and stimulated with HA antigen *ex vivo*, then YA females, and to a lesser extent OA females, had greater numbers and sizes of HA+IgG+ ASCs than their age-matched male counterparts at 7 DPV. These data highlight that the propensity for ASCs to secrete IgG in response to HA antigen, particularly H3, is consistently greater for females than males, which was caused by ER signaling as these effects were inhibited by ER antagonism. FACS-sorted CD19+ B cells from YA females also had greater transcriptional activity at 7 DPV than YA males, with upregulation of estrogen-responsive genes and NF-κB-mediated inflammatory pathways in several B cell subsets, including ASCs and memory B cells.

With YA females having greater concentrations of estrogens than either YA males or OAs of either sex, we wanted to consider how estrogens could impact the activity of B cells. Estrogens have their biological effects primarily by binding to ERs in the cytoplasm of cells, including B cells, to have their canonical effects by serving as transcriptional factors that can up or downregulate the expression of genes that contain EREs. In the present study, DEGs in YA females, particularly at 7 DPV, expressed more EREs in their promoters than did DEGs in either YA males or OAs of either sex. Whether ERE activation causes up- or downregulation of DEGs cannot be determined from our data alone. Several of the DEGs upregulated in YA females, as compared to males, however, have been empirically shown to be upregulated in the presence of estrogens in cell lines (e.g. *LINGO1, GADD45B, TEX14, POU3F1*); whereas DEGs upregulated in YA males (e.g., *BIK*) have been shown to be downregulated in the presence of estrogens (*34*).

In addition to directly binding EREs, cytoplasmic ERs can interact with other transcriptional factors (e.g., NF-κB) to alter gene expression, independent of EREs (*35*). In pseudobulk and single cell analyses, TNF signaling via NF-κB pathways were enriched in several CD19+ B cell subpopulations, including ASCs and memory B cells, of YA females as compared to YA males, with this sex difference being maintained in OAs. Activation of B cells with antigen can upregulate NF-κB signaling (*36*), which could be regulated by estrogenic activation of ERs. NF-κB signaling is critical in the development, survival, and activity of B cells (*37*). TNF signaling in B cells plays roles in activation and differentiation of germinal center B cells as well as antibody production by ASCs. TNF signaling also is critical for upregulating B lymphocyte-induced maturation protein-1 in B cells to promote ASC differentiation (*38*), which might be greater in reproductive aged females than males, based on our data. TNF signaling is critical for production of high affinity antibodies to antigens (*39*), possibly including vaccine. Activation of IL-6 signaling pathways also was greater in YA females than males across naïve, memory, and ASCs. IL-6 activation is downstream of TNF signaling in B cells and is critical for germinal center formation, including in the context of systemic lupus erythematosus (SLE), which occurs at a ratio of 9:1 for females compared with males (*40*). We confirmed that estrogen and inflammatory signaling pathways interact to cause greater vaccine-induced antibody responses in YA females because E2 upregulation of antibody secretion in B cells from YA females required signaling through NF-κB and TNF.

Preclinical animal models provide insights into sex differences and the effects of aging and gonadal steroids on influenza vaccine-induced immunity and protection. Vaccination of adult female mice results in greater quantity and quality of vaccine-specific antibodies, greater class switched and somatic hypermutated antibodies, greater expression of *Tlr7,* and greater numbers of antibody producing B cells in draining lymph nodes (*9, 22, 41, 42*). In adult mice, greater vaccine-induced immunity in females results in better protection than males from infection and severe disease after live virus challenge. Also, like humans, vaccine-induced immunity and protection against influenza is mitigated in old age mice, which is caused by reduced estrogen levels in females. If estrogens are replaced in aged female mice, their vaccine-induced antibody responses are increased to levels observed among young adult females and they are better protected against disease following live influenza virus challenge (*22*). Through use of the transgenic four core genotype mice, where gonadal sex can be decoupled from sex chromosome complement, following influenza vaccination, gonadal females (i.e., mice that have ovaries), regardless of their sex chromosome complement (XX or XY) have greater numbers of germinal center B cells and ASCs, produce higher titers of nAb, and are better protected against live virus challenge than gonadal males (i.e., XX or XY mice that have testes) (*22*). These data highlight that among both humans and mice, age-related sex differences in vaccine-induced immunity are caused by changes in circulating estrogens. Random forest models further illustrate that prior to vaccination, steroids, including 17-hydroxyprogesterone, estrogens, and testosterone, as well as HA+CD19+ B cells and ASCs are major predictors of seroconversion 28 DPV, particularly in YA. This provides additional evidence for gonadal steroid mediation of greater influenza vaccine-induced immunity among reproductive-aged females as compared with age-matched males and explains how sex differences in vaccine-induced immunity are reduced with older age.

Old adults manifest greater frequencies of ABCs and greater enrichment of inflammatory pathways in B cells prior to vaccination in both males and females. ABCs tend to accumulate in aged individuals and during chronic exposure to antigen, including autoantigens (*43*). Aging also is associated with a low-grade, systemic and unsolved chronic inflammatory state termed inflammaging (*44*) or chronic low-grade inflammatory phenotype (*45*), with a significant impact on innate and adaptive immune responses following vaccination (*46*). In addition to greater inflammatory profiles in B cells prior to vaccine, our data suggest that after vaccination, aging is associated with greater IFN signaling in B cell subsets, including ASCs and memory B cells, but only among OA males. In contrast, among YA, females trended toward having greater enrichment of IFN signaling pathways in memory B cells than males. IFN signaling, including type I IFN signaling in B cells has been well characterized in the context of autoreactive B cells. In mice, induction of SLE results in an accumulation of ABCs with upregulation of interferon stimulated genes and *Tlr7* gene duplication (*47*). Also in mice, type I IFN signaling is involved in T cell independent antibody production by B cells (*48*). Much like the pattern of estrogen concentrations, in which concentrations were greater in YA females and OA males as compared with their age-matched counterparts, it appears that IFN signaling activation, particularly in memory B cells, is elevated in these individuals after vaccination. Our previous studies in OAs that received the high dose QIV reveal that estradiol concentrations are greater in males than females and are associated with greater concentrations of IL-6 and reports of adverse events in males compared with females (*49*). Sex differences and effects of aging on TNF and type I IFN pathways have been characterized in the context of developing antibody-mediated autoimmune diseases, e.g., SLE. We now show that these pathways are important in vaccine-induced immunity, which requires further investigation.

There are several limitations to this study. We utilized two cohorts of adults, both with high levels of pre-existing immunity due to high rates of annual influenza vaccination. Our data highlight that sex differences in influenza vaccine-induced immunity are more pronounced in people with less pre-existing immunity (i.e., responders). The cohorts in the current study each received different doses of QIV creating a confound for age-related comparisons of post-vaccine immune responses. A previous comparison between similar YA and OA cohorts in Baltimore, Maryland revealed that in response to the first two doses of COVID-19 mRNA vaccine, the age-related decline in immunity was greater for males than females when OA and YA were compared (*50*). This study also revealed that in the absence of pre-existing immunity to SARS-CoV-2, OA females maintained greater durability of antibody over time than OA males.

Together, these data further highlight that sex differences are observed in de novo antibody responses of OA. How steroids impact antibody responses in older individuals requires further inquiry as it appears that estrogens do not increase antibody secretion by B cells from OA males, but elevated concentrations of estrogens in OA males are associated with enrichment of estrogen response signaling pathways in B cell subsets, similar to YA females. Self-reported BC use did not impact either antibody or cellular measures post vaccination, but small sample sizes precluded deeper analysis into the type of BC used, which ranged from progesterone only oral contraceptives and combined estrogen and progesterone oral contraceptives to non-hormonal intrauterine devices. This study did not enroll adults 50-74 years of age, a group that would encompass menopausal women. The observation that aging of cells is nonlinear with bursts of transcriptional dysregulation occurring at ages including mid 40s and early 60s in humans (*51*) further highlights the need to analyze males with comparisons to females in the postmenopausal period when some females might be taking hormone replacement therapy (HRT). Future studies should consider how menopause and the use of estrogen-based HRT affect vaccine-induced immunity as we are showing a prominent role for estrogen signaling in B cells.

Greater antibody responses among females compared with males, especially among reproductive aged individuals are well conserved across species and diseases (*52*). Whether in response to vaccine antigens, innocuous antigens (e.g., allergens), or self-antigens (e.g., autoimmunity), females mount greater antibody responses than males (*53*). Harnessing sex differences for public health could translate into considering different vaccine formulations based on sex, e.g., developing adjuvants to increase estrogen signaling in B cells in males of diverse adult ages and old females. This could include use of TLR7 adjuvants as *TLR7* is both an X-linked gene that escapes X-inactivation and is more highly expressed in B cells from females than males (*42, 54*). *TLR7* also has EREs in its promoter (*55*). What if estrogens could be administered as an adjuvant with vaccines to boost B cell signaling and antibody secretion, which could be beneficial for postmenopausal women? There is significant heterogeneity in influenza vaccine effectiveness and consideration of age and sex differences in precision vaccinology could lead to better health outcomes for everyone.

## MATERIALS AND METHODS

### Study design

Our study involved two separate cohorts of human participants (young adults, 18-49 years old and old adults, 75+ years old), which were approved by the Johns Hopkins Institutional Review Boards (IRB00091667, NA_00092365). Complete sample sets were collected for 83 young adult healthcare workers (HCWs) who were enrolled through the Johns Hopkins Centers of Excellence for Influenza Research and Response (JHCEIRR) annual vaccination program. Complete sample sets were available for 84 old adults from the Johns Hopkins Longitudinal Influenza Immunization Study of Aging over 75 years of age (JH LIISA 75+). Old adults with a history of allergic reaction to influenza vaccines or to eggs, were currently taking oral steroids or immunosuppressive medications, or had worsening or new-onset of immune-modulating conditions (e.g., rheumatoid arthritis, hematologic malignancies, etc.) were excluded. Written consent was obtained for all participants as part of the enrollment processes. Participant demographics were self-reported. Study data were collected and managed via REDCap electronic data capture tools hosted at Johns Hopkins University (*56, 57*). After the pre-vaccination blood draw (0 DPV), standard dose inactivated quadrivalent influenza vaccines or high-dose inactivated quadrivalent influenza vaccines were administered intramuscularly to young adult and old adult cohorts, respectively. Blood samples were subsequently collected at 7-and 28-days post-vaccination (DPV) and peripheral blood mononuclear cells (PBMCs), and plasma were separated and stored in liquid nitrogen or at -80°C, respectively.

### Influenza vaccine

The QIV formulation consisted of two influenza A and two influenza B strains based WHO recommendations for the Northern Hemisphere for the 2019–2020 season: A/Brisbane/02/2018 (H1N1)pdm09-like virus, A/Kansas/14/2017 (H3N2)-like virus, B/Phuket/3073/2013-like virus (B/Yamagata/16/88 lineage), and B/Colorado/06/2017-like virus (B/Victoria-lineage/2/87 lineage)(*18*).

### Viruses and recombinant HA antigens

The influenza A/Brisbane/02/2018 IVR-190 virus and influenza A/Kansas/14/2017 X-327 virus was obtained from the US Centers of Disease Control and Prevention (CDC). The infectious virus titer was determined on the MDCK-Siat cell using a 50% tissue culture infectious dose (TCID50) assay as previously described (*9, 58*). For the flow cytometry, glycosylated recombinant HA proteins (eEnzyme) were C-terminal 6x His-tagged influenza full-length hemagglutinin (H1N1) (A/Brisbane/02/2018)-pdm09-like (amino acid 18-530) (GISAID accession# EPI1312566) or C-terminal 6x His-tagged influenza hemagglutinin (A/Kansas/14/2017/H3N2) (aa 17-529)(GenBank accession#: AXQ12067). For flow cytometry staining, recombinant HA proteins were biotinylated using the EZ-Link Micro NHS-PEG4-Biotinylation Kit (Thermo Fisher).

### Flow cytometry

Cryopreserved peripheral blood mononuclear cells (PBMCs) were thawed and resuspended in flow washing buffer (FB), composed of phosphate-buffered saline (PBS) supplemented with 1% fetal bovine serum (FBS), 25 mM HEPES, and 1 mM EDTA. Cells were washed twice with PBS and incubated with LIVE/DEAD™ Fixable Aqua Dead Cell Stain dye (Thermo Fisher, #L34957) for 30 minutes at room temperature to assess viability. Live cells were identified based on exclusion of the Aqua dye. The average post-thaw viability of PBMCs was approximately 95%, with an average recovery of 80% of live cells from frozen samples. For single-cell RNA sequencing (scRNA-seq), PBMCs were stained on ice for 30 minutes with a panel of cell surface antibodies (**Table S2**). For HA-specific cells stimulation, PBMCs were incubated on ice for 30 minutes with 2 µg of biotinylated recombinant hemagglutinin (HA) peptide pools specific to H1N1 (A/Brisbane/02/2018, aa 18-530, eEnzyme, #IA-H1-B18WP) and H3N2 (A/Kansas/14/2017, aa 17-529, eEnzyme, #IA-H3-K17WP). Staining was performed using a characterized panel of cell subtype markers (**Table S2**) diluted in Brilliant Staining Buffer (BD Horizon). Following staining, cells were washed twice with FB and resuspended in 200 µL of FB for acquisition on a BD Symphony A3 flow cytometer. For control conditions, unstimulated samples were processed in parallel without HA peptides, and negative control was included using only streptavidin. Fluorescence-minus-one (FMO) controls were used for each fluorophore to assess background and determine gating thresholds. Following stimulation for flow cytometry, cells were processed for downstream analysis, including flow cytometry-based phenotyping and cell sorting. Compensation values were established before acquisition using appropriate single-stain controls. Antibody secreting cells (ASCs) were defined as CD3e^−^CD14^−^ CD16^−^CD19^+^IgD^−^CD71^+^CD38^+^CD20^−^ cells and Memory B (MB) cells were defined as CD3e^−^CD14^−^CD16^−^CD19^+^IgD^−^CD71^−^ as previously described (*20*) (**Fig. S10** and **Table S3**). Age-associated B cells (ABCs) were defined as CD3e^-^CD14^-^CD16^-^CD19^+^CD21^-^CD11c^+^Tbet^+^ (**Table S3**). The flow cytometry results were analyzed using FlowJo™ v10.8 Software (BD Life Sciences).

**Supplementary Materials Materials and Methods**

**Microneutralization assay**

**ELISpot**

**Liquid chromatography mass spectrometry**

**Pseudobulk differential gene expression and gene set enrichment analysis**

**Putative estrogen responsive elements in promotor sequences**

**qRT-PCR**

**Statistical analyses of quantitative data**

**Machine learning algorithm**

**Fig. S1**: Among young, but not old adults, neutralizing antibody (nAb) titers were greater among females than males and were not affected by birth control (BC) usage.

**Fig. S2:** Gating strategy for flow cytometric analyses of CD19+ B cell subsets in peripheral blood mononuclear cells (PBMCs) from young and old adults collected prior to and after receipt of seasonal influenza vaccination.

**Fig. S3:** Among young, but not old, adults, females had greater HA+ B cells than males.

**Fig. S4:** No effect of birth control (BC) use on HA+ B cells among young adult females.

**Fig. S5:** Old adults had greater frequencies of age-associated B cells (ABCs) than young adults. **Fig. S6:** Random forest modeling performance metrics for classifying H3N2 seroconversion status among young and old adults using D0 measures.

**Fig. S7:** Steroidal and immunological profiles of young and old adult males and females from whom CD19+ B cells were sorted and used for single cell RNA sequencing (scRNAseq).

**Fig. S8:** Identification of B cell subsets from single cell RNA sequencing of FACS-sorted CD19+ B cells in peripheral mononuclear blood cells (PBMCs) collected from young and old adults.

**Fig. S9:** Prior to vaccination, CD19+ B cells from young and old adults had distinct transcriptional profiles among male and female participants.

**Fig. S10:** Estrogen receptor signaling enhanced H3N2-, and to a lesser extent H1N1-specific IgG production in female antibody-secreting cells compared to males, independent of age.

**Table S1:** Summary of PBMC viability following ELISpot inhibitor treatments

**Table S2**: qPCR Primer sequences

**Table S3:** Flow cytometric primary antibodies used for characterizing human B cell subsets

**Table S4:** Cell markers of human total and HA-specific CD19+ B cell subsets used for flow cytometric analyses

## Data Availability

All code can be accessed through GitHub at https://github.com/ayin0510/Influenza-Vaccine-Human-B-Cell-Project-2019-2020. The scRNA-seq transcriptomic data generated in this study have been deposited to the NIH GEO database (https://www.ncbi.nlm.nih.gov/geo/). All data are available in the main text or the supplementary materials.

https://www.ncbi.nlm.nih.gov/geo/

## Acknowledgments

We thank the clinical coordinators, nurses, and staff who assisted with sample collection. We thank Sabal Chaulagain, Janna Shapiro, Nico Swanson, and Caelan Barranta for assay assistance. We thank Ali Ellebedy and his lab for sharing flow cytometry protocols and insights for identifying influenza-specific antibody secreting cells. We thank Jacquelien J.G. Hillebrand from the Endocrine Laboratory at the University of Amsterdam, The Netherlands for conducting estrone and estradiol LC-MS/MS. We thank Engle Abrams, Yan Chen, and Guodong Zhu for assistance with the OA cohort. We thank the members of the Klein, Davis, Pekosz, and Thompson labs for detailed discussions about these data.

## Funding

Funding provided the National Institutes of Health NIA/ORWH Johns Hopkins Specialized Center of Research Excellence in sex differences (U54AG062333; SLK) and the NIAID Johns Hopkins Center of Excellence for Influenza Research and Response (75N93021C00045; AP and RR), with additional support from the Paul and Irma Milstein Foundation Program at the Johns Hopkins Center on Aging and Immune Remodeling (SXL).

## Author contributions

Conceptualization: SLK, RWM, PG, SXL, AP

Methodology: HSP, AY, HL, AJ, AD, PG, HJ, WZ, EFEW, SAAVDB

Investigation: KF, HL, JSL, RER

Visualization: HSP, AY, WZ, EFEW, AW

Funding acquisition: SLK, SXL, AP, RER

Project administration: PJS, KZJF, RWM, SLK, RER, SXL, AP

Supervision: SLK, PG, HJ, SXL, AP

Writing – original draft: HSP, AY, SLK

Writing – review & editing: all authors

## Competing interests

SXL received in-kind support from Sanofi, Inc, including influenza vaccine (Fluzone® High Dose) for the OA cohort.

## Data and materials availability

All code can be accessed through GitHub at https://github.com/ayin0510/Influenza-Vaccine-Human-B-Cell-Project-2019-2020. The scRNA-seq transcriptomic data generated in this study have been deposited to the NIH dbGaP database (https://dbgap.ncbi.nlm.nih.gov/home). All data are available in the main text or the supplementary materials.

## Notes

### Competing Interest Statement

The authors have declared no competing interest.

### Author Declarations

Our study involved two separate cohorts of human participants (young adults, 18-49 years old and old adults, 75+ years old), which were given ethical approval by the Johns Hopkins Institutional Review Boards (IRB00091667, NA_00092365).

### Summary of Updates

Additional experiments were added, including evaluate changes in the expression of genes with estrogen response elements along the TNF signaling via NF-kB mediated pathway in the presence or absence of estradiol ex vivo. Additional experiments were added, including ex vivo analyses of estradiol-induced IgG secretion by antibody secreting cells in the presence or absence of NF-kB or TNF inhibitors.

