## Supplementary material for "Estrogen upregulates NF-κB and TNF signaling in B cells to cause sex and age differences in antibody responses to seasonal influenza vaccination": Methods and Supplemental Figures and Tables

**Supplementary Materials:**
**Supplementary Materials and Methods**

### Microneutralization assay

Serum samples were diluted with receptor-destroying enzyme (RDE, Denka Seiken) at 1:3 ratio and incubated overnight at 37 °C followed by heat inactivation at 56 °C for 30 min. Samples were 2-fold serially diluted in Incomplete media (IM), composed of DMEM with 1%P/S and L-Glutamine (Incomplete medium, IM) mixed with 100 TCID50 of each virus (A/Brisbane/02/2018 IVR-190 (H1N1) or A/Kansas/14/2017 X-327 (H3N2)), and incubated at room temperature for 1 h. The virus/serum mixture was transferred in duplicate into the 96-well cell culture plates containing confluent SIAT1(cDNA of human 2,6-sialtransferase) stable Expressing Madin-Darby Canine Kidney (MDCK-SIAT1) cells and incubated at 32 °C. After a 24 h incubation, plates were washed once with 1X PBS, fresh IM was added, and cells were incubated for 6 days. Plates were fixed with 4% formaldehyde, stained with naphthol blue-black solution, and scored as described previously (*1, 2*).

### ELISpot

Direct *ex-vivo* ELISpot was performed to determine the number of total or recombinant HA-binding IgG-secreting cells present in PBMC samples. ELISpot plates (#MSIPS4W10, Millipore) were activate membrane with 35% EtOH and wash with PBS. After plate was coated overnight at 4°C with 50ul/well of carbonate buffer (pH 9.6) containing 3 μg/mL recombinant HA proteins (#IA-H1-B18WP (H1) and #IA-H3-K17WP (H3), eEnzyme). Plates were blocked the following morning for 90 min at 37°C with no phenol red-RPMI 1640 supplemented with 10% FBS charcoal stripped serum (#12676029, ThermoFisher). For HA-specific IgG, dilutions of washed PBMCs were incubated for 18 h in no phenol red-RPMI supplemented with 10% FBS charcoal stripped serum, 2mM L-glutamine (# A29168, Gibco), 100U/mL Penicillin-Streptomycin (#15140-122, Gibco), at 37 °C. To stimulate B cells with estradiol (E2), PBMC were cultured in complete media for 1h containing RPMI 1640 medium (MilliporeSigma) with 10% FBS charcoal stripped serum (#12676029, Gibco), 2mM L-glutamine (# A29168, Gibco), 100U/mL Penicillin-Streptomycin ( #15140-122, Gibco), 7.5 mM HEPES (#15630, Gibco), 1 mM sodium pyruvate (#11360-070, Gibco), essential amino acid solution (#11130-051, Gibco), nonessential amino acid solution (#11140-050, Gibco) at 37°C (*3, 4*). Cells were incubated in Tamoxifen (1uM, #J63509.ME, ThermoFisher Scientific) or TNF inhibitor (Etanercept, 1ug/mL , # HY-108847, MedChemExpress) or NF-kB inhibitor (Bay 11-7082, 1uM, #196870, Sigma) in complete media 1 hour before E2 treatment at 37°C. An hour later, E2 (100,000 pmol/L (=$1*10^{-7}$ mol/L, 100nM #3301, Millipore) was added to select wells in complete media for 18h at 37°C. After washing the plates, secreted antibodies were detected with anti-human IgG-biotin (#2048-08, 1:2000, Southern biotech), and plates were incubated at 37°C for 2 hours. Plates were washed 3 times with PBS before adding alkaline phosphatase-conjugated streptavidin (#7105-04, 1:1000, Southern Biotech), and incubated at 37°C for 1h. Plates were washed 5 times with PBS and 3 times with distilled water before adding 5-bromo-4-chloro-3-indolyl phosphate (BCIP)/nitro blue tetrazolium (NBT) 1-step solution (Thermo Fisher Scientific, #34042) (50 μL/well) and incubating at 37°C for 15–20 minutes. Development of the BCIP/NBT substrate was terminated by decanting the plates and washing 3 times with distilled water. Plates were read on an

Immunospot plate reader (Cellular Technology). Data were analyzed using Immunospot version 3.0 software. Data are presented as ASCs per $1 \times 10^6$ cells.

**Selection and dosing of inflammatory inhibitors**

We treated PBMCs collected at 7 DPV from YA females with either media alone (untreated), E2 alone, or pre-treated with TNF or NFκB inhibitors prior to E2. The TNF inhibitor used for these experiments, Etanercept, was chosen among several anti-TNF monoclonal antibodies that have been FDA-approved for treatment of autoimmune conditions. Etanercept inhibits TNF from binding to the TNF receptor, thereby preventing signaling via the NFκB or MAPK pathways. The NFκB inhibitor selected, BAY 11-7082, is an agent that inhibits the phosphorylation of IκBα induced by TNF, thereby inhibiting the signaling of NFκB via TNF. The TNF inhibitor Etanercept was used at 1 μg/mL, based on in vitro studies showing effective inhibition of TNF-induced signaling in human $CD4^+$ T cells without affecting cell viability(*5*). The NFκB inhibitor BAY 11-7082 was used at 1 μM, consistent with previous reports demonstrating effective NFκB inhibition in human PBMCs and plasmacytoid dendritic cells while maintaining cell viability(*6*). We tested these inhibitors at multiple doses to evaluate effects on cell viability and antibody secretion (**Table S1**).

**Liquid chromatography mass spectrometry**

Sex hormones were measured using liquid chromatography mass spectrometry under ISO15189 accreditation. Estrogens (estrone [E1] and estradiol [E2]) were measured at the Endocrine Laboratory in Amsterdam UMC, Amsterdam, The Netherlands using a well-defined high performance liquid chromatography, electrospray ionization MS/MS (UPLC-ESI-MS/MS) described (*7*). Estradiol and estrone (Cerilliant CRM) and 13C3 labelled estradiol (Isosciences) and estrone (Sigma Aldrich) were used as standards and internal standards respectively. All samples were run in duplicate. Testosterone was measured at the Diagnostic Laboratory of Endocrinology in Erasmus MC, Rotterdam, The Netherlands, in a full steroid profile also including precursors, metabolites, and other steroids including androstenedione, dehydroepiandrosterone (DHEA), dehydroepiandrosterone sulfate (DHEAS), 11-desoxycortisol, corticosterone, cortisol, 21-deoxycortisol, 17-OH-progesterone and dihydrotestosterone (DHT). Erasmus MC uses a well-defined and highly sensitive high performance liquid chromatography, electrospray ionization MS/MS (UPLC-ESI-MS/MS) assays which is ISO15189 accredited. Where applicable, the assay has been standardized to reference measurement procedures (e.g. testosterone is a CDC HoSt certified assay). Metabolites are organically extracted from 20-100 μL of serum after protein precipitation. Heavy-labeled internal standards are combined with each sample to enable isotope dilution mass spectrometry and injected onto the mass spectrometer for analysis. Instrumentation used for these analyses is an IVD Waters XEVO TQ-XS UPLC/MS-MS solution. For the metabolites: sera are purified by solid phase extraction, and chromatographic separation is performed using a reversed phase chromatography (12-minute gradient). Injection into the mass spectrometer utilizes electrospray ionization in both positive and negative modes. For all hormone analyses for quality control, a systems suitability quality control is performed, as well as three level matrix matched control sera internal quality control program, using the same techniques described above. Quality control programs are run with each run. External quality assessment is performed at least 6 times per calendar year to ensure proper calibration of the entire instrumentation. For metabolites that are included in the Joint Committee for Traceability in Laboratory Medicine, trueness has been verified. Results of all samples are determined by response, relative retention time, raw signal, peak shape, ion ratio, signal/noise ratio and chromatographic resolution. Chromatographic integration is performed by Waters MassLynx software.

**Single cell RNA-sequencing (scRNA-seq) library prep and data processing**
scRNA-sequencing was performed on n=4/sex/timepoint for each cohort from the 2019-2020 season. Sorting was performed on a BD Melody cell sorter, and cells were collected into 1.5 mL Eppendorf tubes containing PBS with 10% FBS. FACS-sorted CD19+ B cells (7-AAD-CD3-CD14-CD16-CD19+) in PBS with 10% FBS were immediately processed through 10X Chromium. The cells were gently pelleted, and volume reduced prior to counting and assessing viability on the Countess 3 (ThermoFisher), according to manufacturer's protocol. The cell suspension volume calculator table (10x Genomics) was used to determine targeted cell recovery for each sample based on cell counts. A volume targeting approximately 10,000 cells was immediately processed following the 10x Genomics' Chromium Next Gem Single Cell 5' Reagents kits v2 (Dual Index) User Guide protocol. Yield and quality of cDNA and 5' GEM libraries were assessed by High Sensitivity D5000 ScreenTape on the 4200 TapeStation (Agilent Technologies). Final libraries were quantified with Qubit High Sensitivity DNA assay on the Qubit Flex Fluorometer (ThermoFisher). Libraries were diluted and equimolar pools prepared, according to manufacturer's protocol for appropriate sequencer. An Illumina iSeq Sequencer with iSeq100 i1 reagent V2 300 cycle kit was used for the final quality assessment of the library pools. For deep sequencing, two 300 cycle (2 x 150 bp) Illumina NovaSeq X Plus (4 lanes, 25 billion reads) runs were performed (Psomagen, Inc). Raw scRNA-seq data were aligned to the human reference genome hg38 (GRCh38.p14) using 10X Genomics Cell Ranger Software. Among YA samples, n=1 female 7 DPV sample and n=1 female 28 DPV sample were excluded from subsequent analyses due to low quality (both have low fraction reads in cells). Among OA samples, n=1 female 0 DPV sample and n=1 female 28 DPV sample were excluded from subsequent analyses due to low quality (one with low number of detected cells and the other with low number of genes detected per cell). Samples within each cohort (i.e., YA and OA) were integrated using Harmony(*8*). B cell subpopulations from scRNA-seq of FACS-sorted CD19+ B cells were identified using Seurat v5 (*9*) based on Louvain clustering of single cells at a resolution of 0.3 and manually annotated using previously reported canonical markers (*10*) for each cohort, respectively.

**Pseudobulk differential gene expression and gene set enrichment analysis**
For pseudobulk analysis of gene expression data, single cell data were aggregated by sample IDs and/or cell clusters. Bioconductor DESeq2 was used to conduct sex by time pseudobulk differential gene expression analysis. Multiple testing was corrected based on the Benjamini-Hochberg procedure to report false discovery rate (FDR) and FDR<0.05 is used to obtain significantly differential genes. The DESeq2 model used to compare sex by time changes within each age group was ~ Sex + Sex:Timepoint + Sex:ind.n to obtain differentially expressed genes (DEGs) between males and females, where ind.n is the nested labeling for samples at 0, 7, and 28 DPV. For age group comparisons at 0 DPV within each sex, the DESeq2 model ~ cohort was used.

Principal component analysis (PCA) was conducted using the *runPCA* function in Seurat v5 using normalized gene count data. DEGs with log2 fold change (log2FC) >|0.5| and padj<0.05

were visualized using 'EnhancedVolcano' (*11*). Hallmark gene set enrichment analysis (GSEA) (*12*)was conducted using all DEGs ranked by log2FC using 'fgsea' following conversion of gene symbols to human Entez gene identifiers in org.Hs.eg.db. Leading edge genes (padj<0.05) for each hallmark gene set were obtained following GSEA and visualized using box plots. Venn diagrams of DEGs (padj<0.05) between young and old adults for each sex were visualized using 'VennDiagram'(*13*).

Single sample gene set enrichment analysis (ssGSEA) was conducted using the 'escape' package (*14*) to calculate hallmark gene set enrichment scores (ES) on a per cell basis across cell clusters at 7 DPV. Mean z-scaled enrichment scores were visualized by heatmaps across the hallmark signaling, immune, and metabolic gene sets. Differential enrichment analysis (DEA) was performed using the *FindMarkers* function on enrichment scores normalized by the nFeature_RNA values for each cell with an FDR=0.05. Normalized enrichment score (NES) differences between males and females for the hallmark TNF signaling via NF-κB gene set were visualized with violin plots, showing the mean and one standard deviation for each sex. All gene expression and gene set enrichment analyses were conducted in R Statistical Software (v4.4.0; R Core Team 2024) either locally or through the Joint High Performance Computing Exchange (JHPCE) in the Department of Biostatistics at Johns Hopkins University.

**Putative estrogen responsive elements in promotor sequences**

To explore whether the vaccine-induced differentially expressed genes could have been regulated by estrogens, corresponding promotor regions of these genes were searched for putative estrogen responsive elements (EREs). The gene of interest was selected in the Human Reference Genome (GRCh38/hg38) to obtain corresponding non-coding promotor region sequences, using a promotor region of -5000 base pairs (TSS) to +3000 base pairs (TSS). EREfinder(*15*), an automated software program, was used to scan sequence data for putative ERE sequences (chosen settings: window size 1000 base pairs, sliding interval 500 base pairs)(*16*). Both palindromic ("perfect") ERE sequence (AGGTCAnnnTGACCT) and imperfect ERE sequences with basepair substitutions were identified, based on predicted binding affinity ($K_d$) to ER α or β using the empirically validated formula of Tyulmenkov and Klinge (*17*). This formula takes into account the important role of (loss of) a perfect half site in ER binding, next to base pair substitutions. A cut-off of $K_d^{-1} > 0.15$ (ERα) was used for identification EREs, which means that identified ERE consist of at least one perfect half site, and the other half site is allowed to have one base pair substitution. Genes with EREs were also manually checked against the EstroGENE database (*18*).

**qRT-PCR of genes with estrogen response elements (EREs)**

PBMCs collected at 7 DPV from YA females were treated for 18 h at 37 °C with either media alone (untreated), E2 alone, or Tamoxifen added 1 h prior to E2 treatment. B cells were isolated using the EasySep™ Human Pan-B Cell Enrichment Kit (#19554, STEMCELL Technologies). Prior to RNA extraction, cell purity was assessed and confirmed to be approximately 95% B cells. Total RNA was extracted using the NucleoSpin XS kit (#740902.50, Macherey-Nagel) according to the manufacturer's protocol, and RNA quantity was measured using the Qubit RNA quantification system. cDNA was synthesized from purified RNA and quantitative PCR (qPCR) was performed on a QuantStudio 6 Pro system (Applied Biosystems) using SYBR Green chemistry in a 96-well plate format. β-actin (ACTB) was used as the housekeeping gene for normalization of gene expression. Primer sequences for all genes analyzed are provided in **Table S4**.

**Statistical analyses of quantitative data**

Live virus neutralization (nAb) titers and sex steroid concentrations were log-transformed and flow cytometric frequencies (%) were arcsine-transformed prior to statistical analyses. Unless stated otherwise, statistical analyses were conducted for YA and OA independently. Linear mixed effects regression models with an interaction term for sex, timepoint, and H3N2 seroconversion status or sex and timepoint were used, adjusting for continuous age. Mann-Whitney U tests, also known as Wilcoxon Rank Sum test, were used to compare ABC+ frequencies between YA and OA or steroid concentration between BC status among YA at 0 DPV. Kruskal-Wallis testing with Benjamini-Hochberg post-hoc corrections at FDR=0.05 were used to compare steroid concentrations at 0 DPV across age groups and sex. All data cleaning and statistical analyses were conducted in Stata 17.0.

**Machine learning algorithm**

A random forest model was trained based on 5-fold cross-validation with 5 repeats to predict seroconversion status from complete baseline sex steroid concentrations, neutralizing antibody, total and HA+ CD19+ flow data of YA and OA. Data were first downsampled to the minor class and then partitioned with 70% used for model training and 30% reserved for model testing. Models were first tuned for mtry values 1 through 10, with the best tuning parameters used with the testing data to classify seroconversion. This process was repeated across 10 different seeds and performance metrics were averaged across the 10 seeds to account for randomness and variability in model performance. Downsampling, random forest modeling, and variable importance analysis were performed using the 'caret' package in R (*19*). Model performance metrics were assessed using 'MLeval'(*19, 20*), 'pROC', and 'caret' packages in R Statistical Software (v4.4.0; R Core Team 2024).

**Figure S1: Among young, but not old adults, neutralizing antibody (nAb) titers were greater among females than males and were not affected by birth control (BC) usage.** In response to H1N1 (**A, C**), all young and old adults, regardless of sex, had H1N1 nAb titers that were above the seroprotection cutoff (nAb titers ≥ 1:40) at 0- and 28-days post vaccination (DPV). In response to H3N2, (**B, D**) 28% of male and 63% of female young adult participants and 44% of male and 37% of female old adult participants were below the seroprotection cutoff prior to vaccination (0 DPV) for H3N2 nAb titers. (**E**) H1N1 and (**F**) H3N2 nAb titers were measure at 0- and 28-days post-vaccination (DPV) among young adult females who were responders (R; nAb ratio≥4) based on their reports of birth control (BC) use. Data collected across multiple timepoints were analyzed by linear mixed effects regression models with an interaction term for birth control use and timepoint, adjusting for continuous age. * represents $p<0.05$ by sex and † represents $p<0.05$ for nAb titers over time.

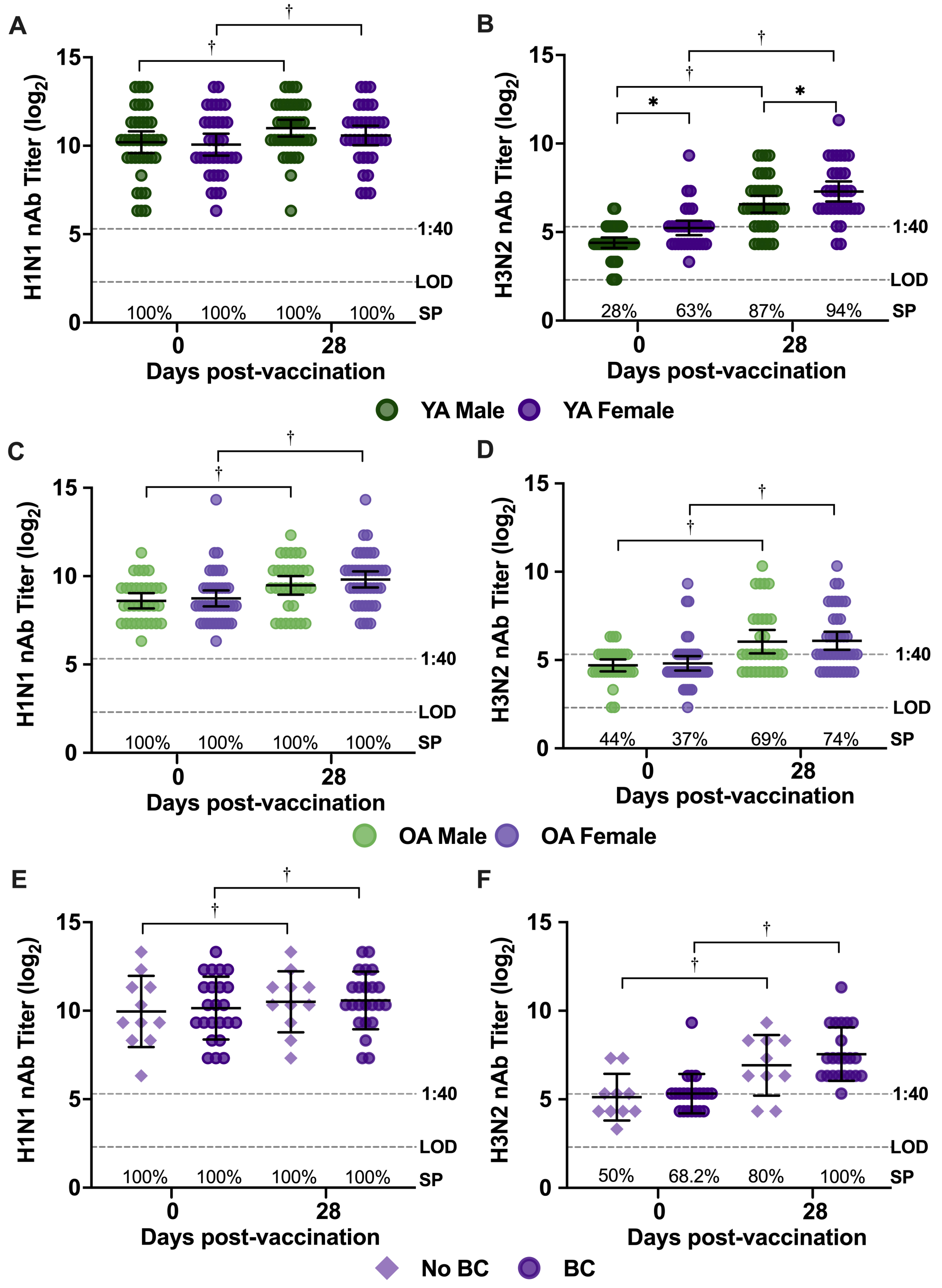

**Figure S2: Gating strategy for flow cytometric analyses of CD19+ B cell subsets in peripheral blood mononuclear cells (PBMCs) from young and old adults collected prior to and after receipt of seasonal influenza vaccination. (A)** Flow gating strategy for memory B (MB; CD3e$^-$CD14$^-$CD16$^-$CD19$^+$IgD$^-$CD71$^-$) cells and antibody-secreting cells (ASCs; CD3e$^-$CD14$^-$ CD16$^-$CD19$^+$IgD$^-$CD71$^+$CD38$^+$CD20$^-$). **(B)** Flow gating strategy for age-associated B cells (ABCs; CD3e$^-$CD14$^-$CD16$^-$CD19$^+$CD21$^-$CD11c$^+$Tbet$^+$).

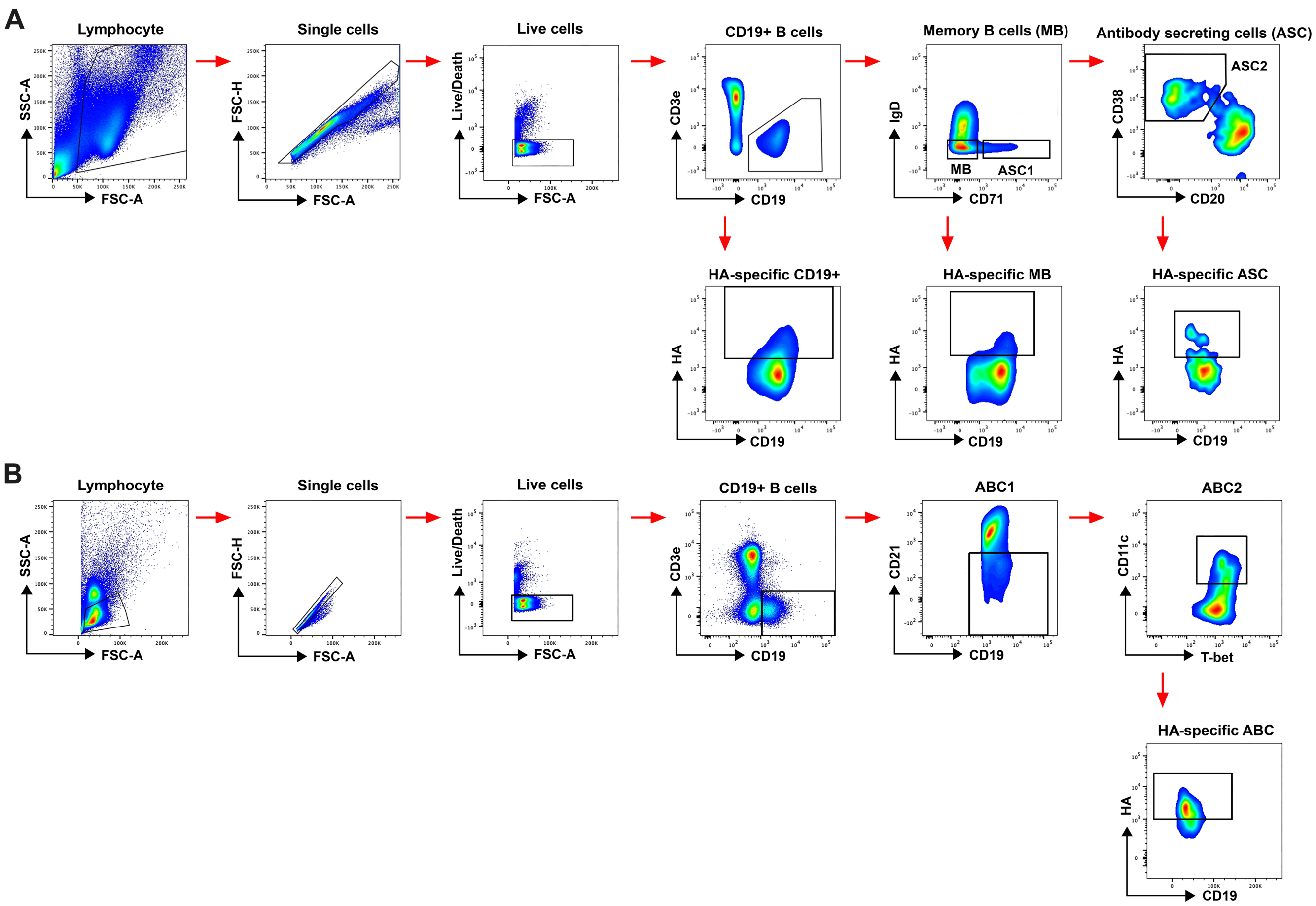

**Figure S3. Among young, but not old, adults, females had greater HA+ B cells than males.** Total and HA+ B cell populations were analyzed by flow cytometry from peripheral blood mononuclear cells (PBMC) collected at 0-, 7-, and 28-days post-vaccination (DPV). Total and HA+ CD19+ B cell frequencies in young (**A-B**) and old (**C-D**) adults. Total and HA+ memory B (MB) cell frequencies in young (**E-F**) and old (**G-H**) adults. Total and HA+ antibody-secreting cell (ASC) frequencies in young (**I-J**) and old (**K-L**) adults. Data collected across multiple timepoints were analyzed by linear mixed effects regression models with an interaction term for sex and timepoint, adjusting for continuous age. Young and old adults were analyzed independently unless specified. ABC data were compared between young and old adults by the Mann-Whitney U test. Dashed lines represent the limit of detection (LOD) and $\log_2$ (1:40 titer) for seroprotection. * represents sex differences with $p<0.05$.

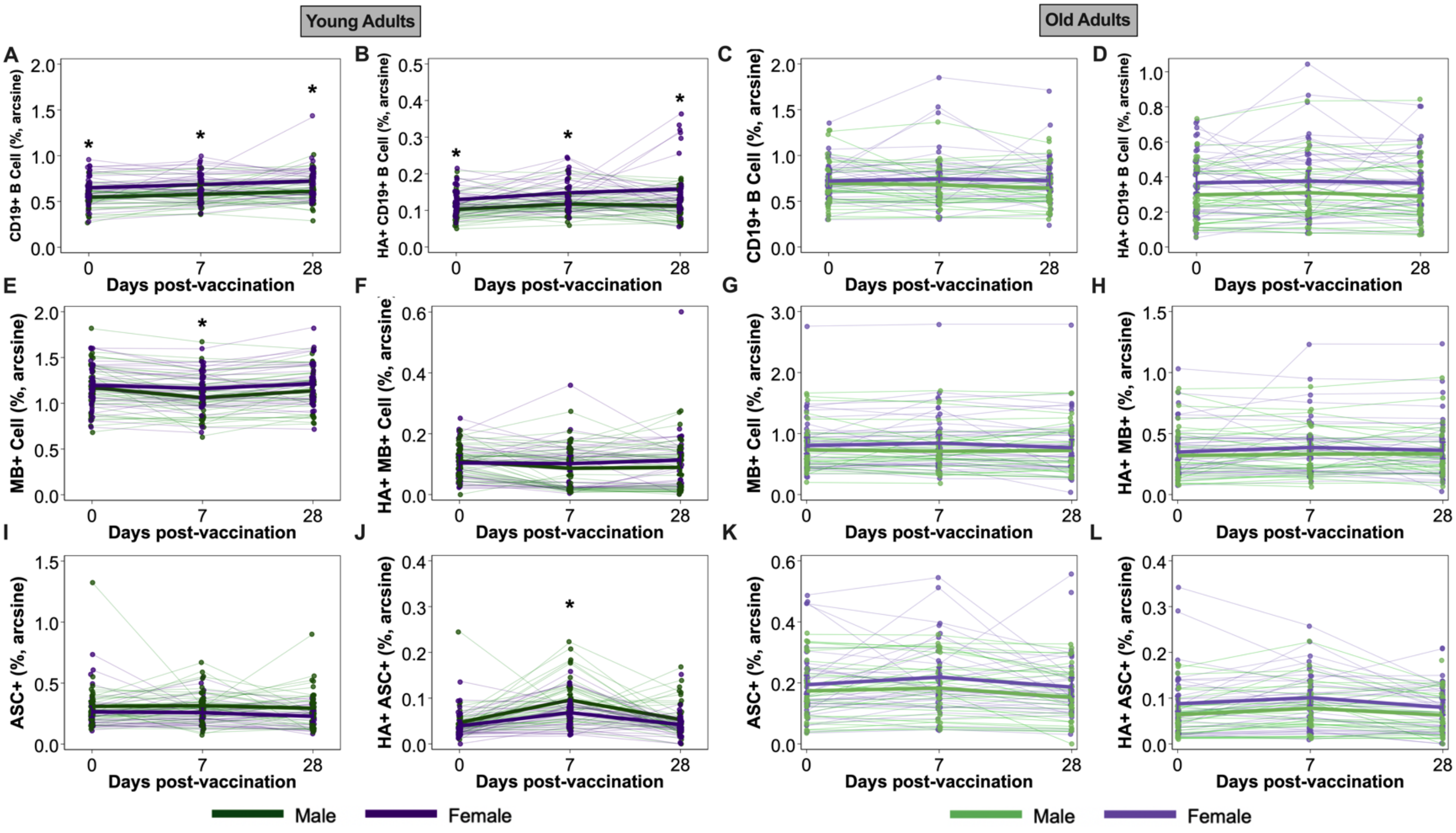

**Figure S4: No effect of birth control (BC) use on HA+ B cells among young adult females.** Total and HA+ B cell populations were analyzed by flow cytometry from peripheral blood mononuclear cells (PBMC) collected at 0-, 7-, and 28-days post-vaccination (DPV). Total and HA+ CD19+ B cell frequencies (**A-B**), total and HA+ memory B (MB) cell frequencies in young (**C-D**) and total and HA+ antibody-secreting cell (ASC) frequencies (**E-F**) were compared between females using birth control (BC) or not using BC. Data collected across multiple timepoints were analyzed by linear mixed effects regression models with an interaction term for sex and timepoint, adjusting for continuous age. Dashed lines represent the limit of detection (LOD) and $log_2$ (1:40 titer) for seroprotection. * represents differences based on hormone-based BC use with $p<0.05$.

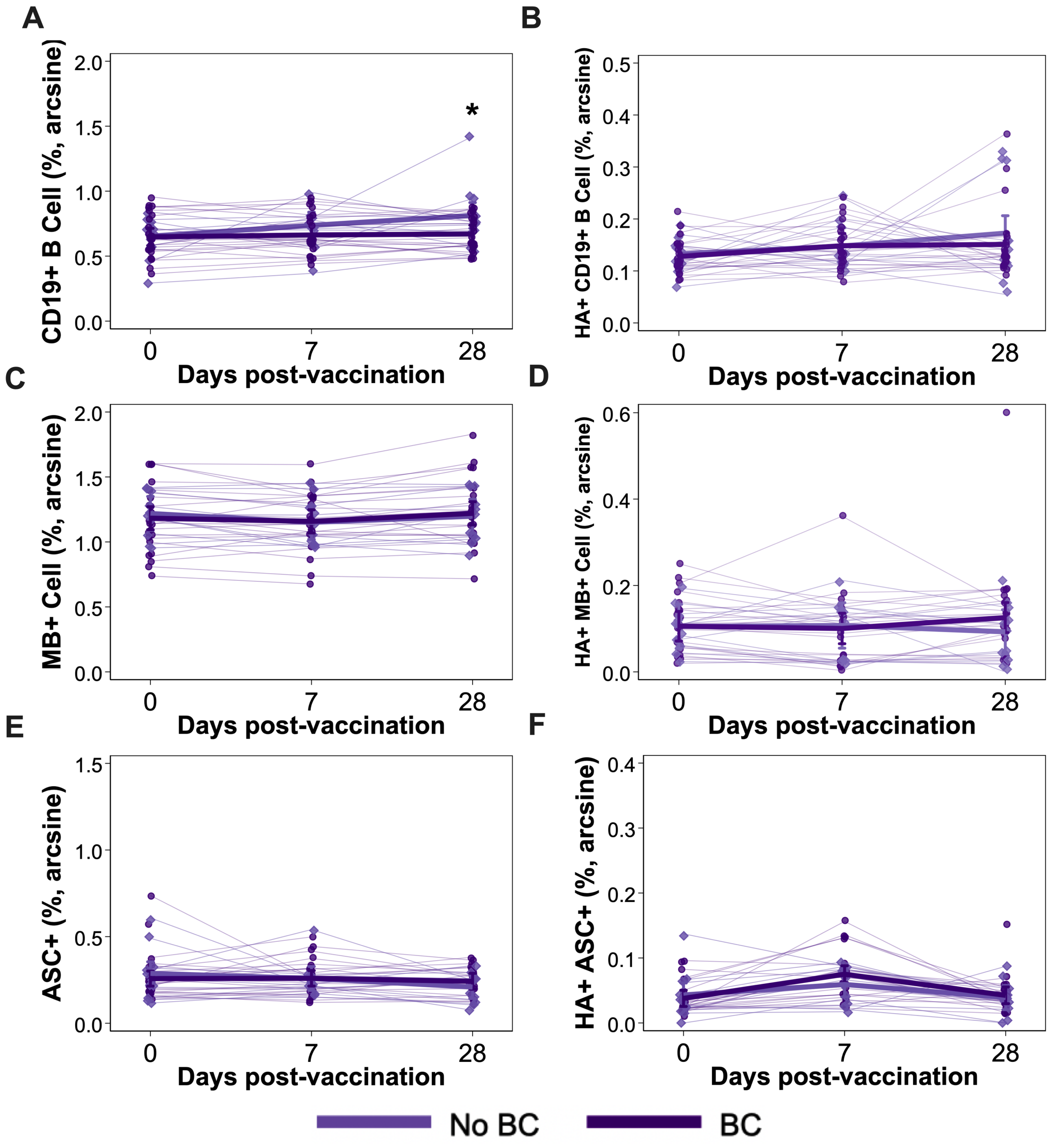

**Figure S5: Old adults had greater frequencies of age-associated B cells (ABCs) than young adults.** B cell populations were analyzed by flow cytometry from peripheral blood mononuclear cells (PBMC) collected at 0-, 7-, and 28-days post-vaccination (DPV). Total and HA+ ABC frequencies (**A-B**) prior to vaccination were compared between a small subset of young adult (n=4) and old adult (n=77) participants. Among old adult males and females, total and HA+ ABC frequencies (**C-D**) were monitored prior to and 7 and 28 days post vaccination. Data collected across multiple timepoints were analyzed by linear mixed effects regression models with an interaction term for sex and timepoint, adjusting for continuous age. ABC data were compared between young and old adults by the Mann-Whitney U test. Dashed lines represent the limit of detection (LOD) and $\log_2$ (1:40 titer) for seroprotection. * represents differences based on age with $p<0.05$.

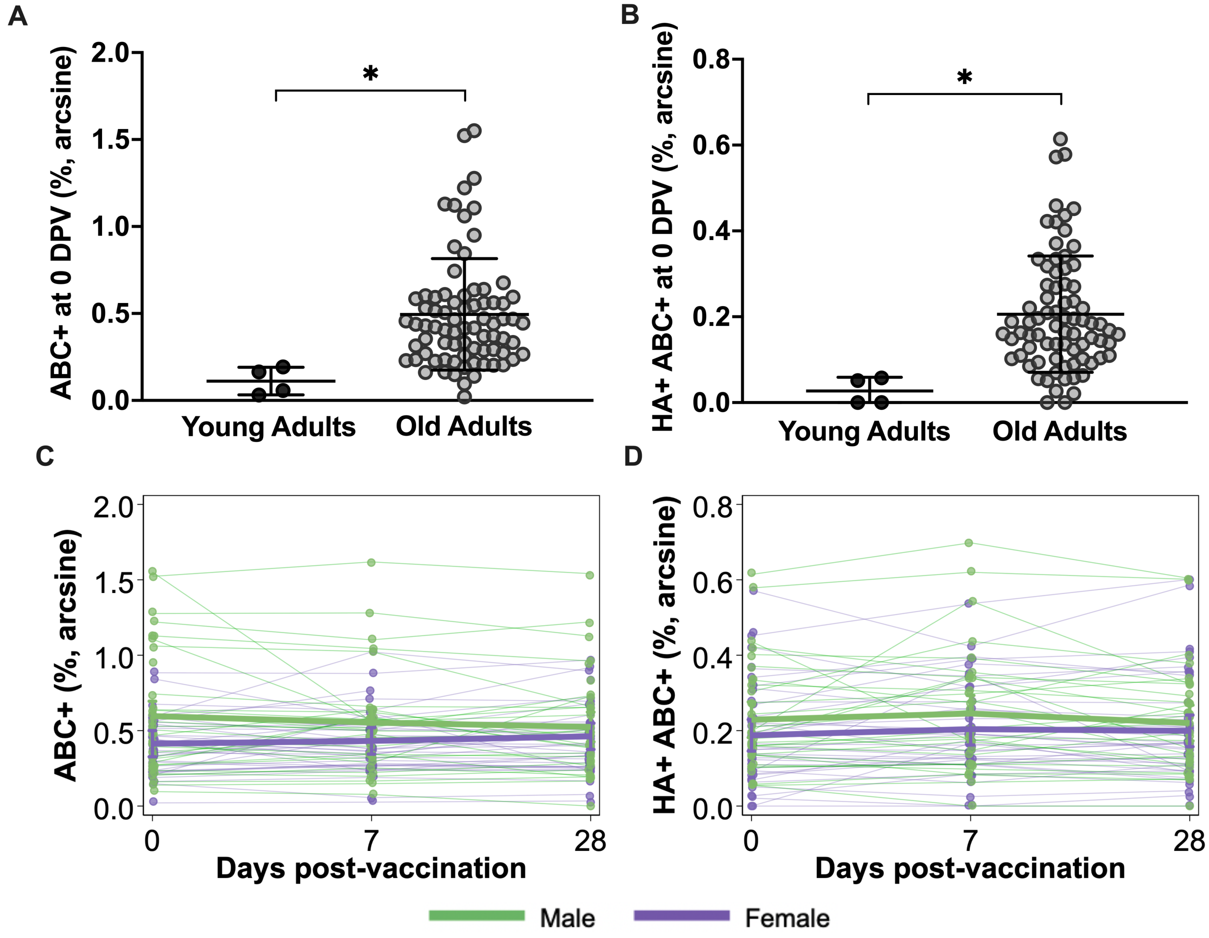

**Figure S6: Random forest modeling performance metrics for classifying H3N2 seroconversion status among young and old adults using D0 measures. (A)** Random forest model sample sizes and performance metrics for all aforementioned models, including area under the receiver operator curve (AUROC), area under the precision-recall curve (AUPRC), and accuracy on the test sets. (**B**) Random forest model sample sizes and performance metrics for the permuted models, including area under the receiver operator curve (AUROC), area under the precision-recall curve (AUPRC), and accuracy on the test sets. All metrics were averaged across 10 distinct seeds. Receiver operator curves (ROC) and precision-recall curves (PRC) of the random forest models classifying by seroconversion status, colored by each seed, for young adult testing set data (**C**). ROC and PRC curves of the random forest models classifying by seroconversion status, colored by each seed, for old adult testing set data (**D**). Receiver operator curves (ROC) and precision-recall curves (PRC) of the random forest models classifying by permuted seroconversion status (i.e., control models), colored by each seed, for the young and old adult training and testing sets (**E-F**), respectively.

**A**

| **Downsampled 5-Fold Cross-Validated Random Forest Model Metrics-Averaged 10x**<br>Predicting participant H3N2 Seroconversion status with 0 DPV (baseline) data | | | | | | |
|---|---|---|---|---|---|---|
| **Model outcome** | **Dataset** | **$N_1$** | **$N_2$** | **AUROC** | **AUPRC** | **Accuracy** |
| | | *Non-Responder* | *Responder* | | | |
| **Seroconversion** | Young Adults | 27 | 27 | 0.61 | 0.54 | 60.63% |
| | Old Adults | 19 | 19 | 0.48 | 0.46 | 54% |

**B**

| **Downsampled 5-Fold Cross-Validated Random Forest Model Metrics-Averaged 10x Control**<br>Predicting participant H3N2 Seroconversion status with 0 DPV (baseline) data | | | | | | |
|---|---|---|---|---|---|---|
| **Model outcome** | **Dataset** | **$N_1$** | **$N_2$** | **AUROC** | **AUPRC** | **Accuracy** |
| | | *Non-Responder* | *Responder* | | | |
| **Permuted Seroconversion** | Young Adults | 27 | 27 | 0.48 | 0.47 | 50.63% |
| | Old Adults | 19 | 19 | 0.38 | 0.38 | 45% |

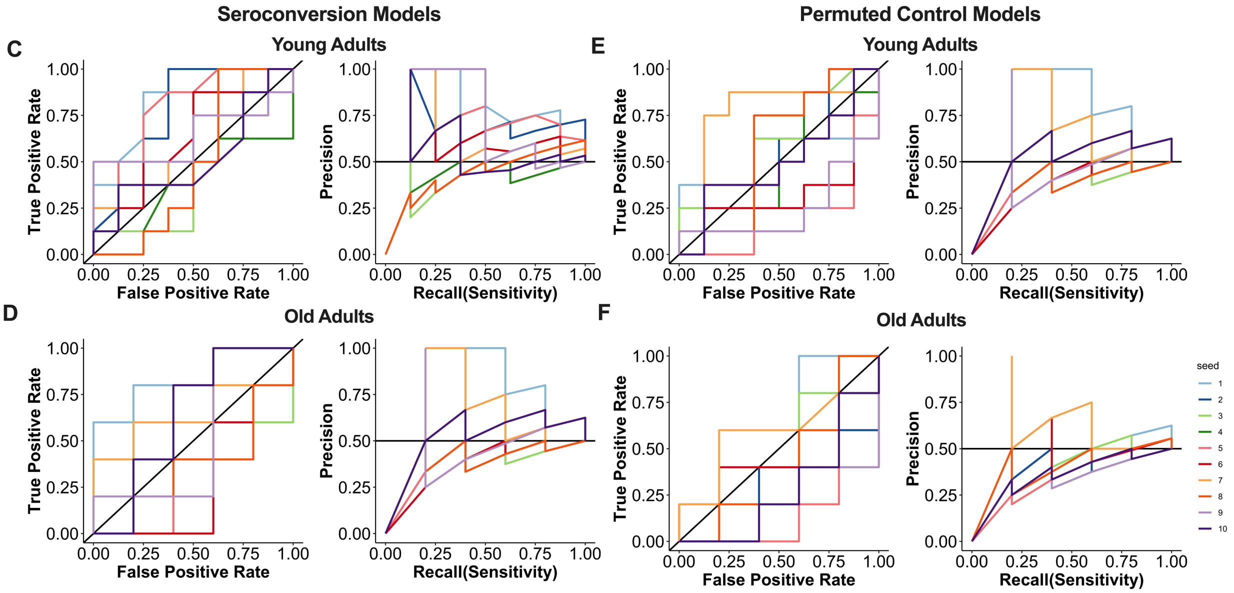

**Figure S7: Steroidal and immunological profiles of young and old adult males and females from whom CD19+ B cells were sorted and used for single cell RNA sequencing (scRNAseq). (A-I)** Baseline concentrations (umol/L, nmol/L, or pmol/L) of steroids as well as their precursors and metabolites, measured from peripheral blood samples collected prior to vaccination (0 DPV). (**J-K, M-N**) Total and HA-specific CD19+ B cell frequencies for young adult and old adult PBMC samples selected for scRNA-seq. (**L, O**) H3N2 neutralizing antibody (nAb) titers at 0 and 28 DPV from young and old adult participants from whom FACS sorted CD19+ B cells were selected for scRNA-seq.

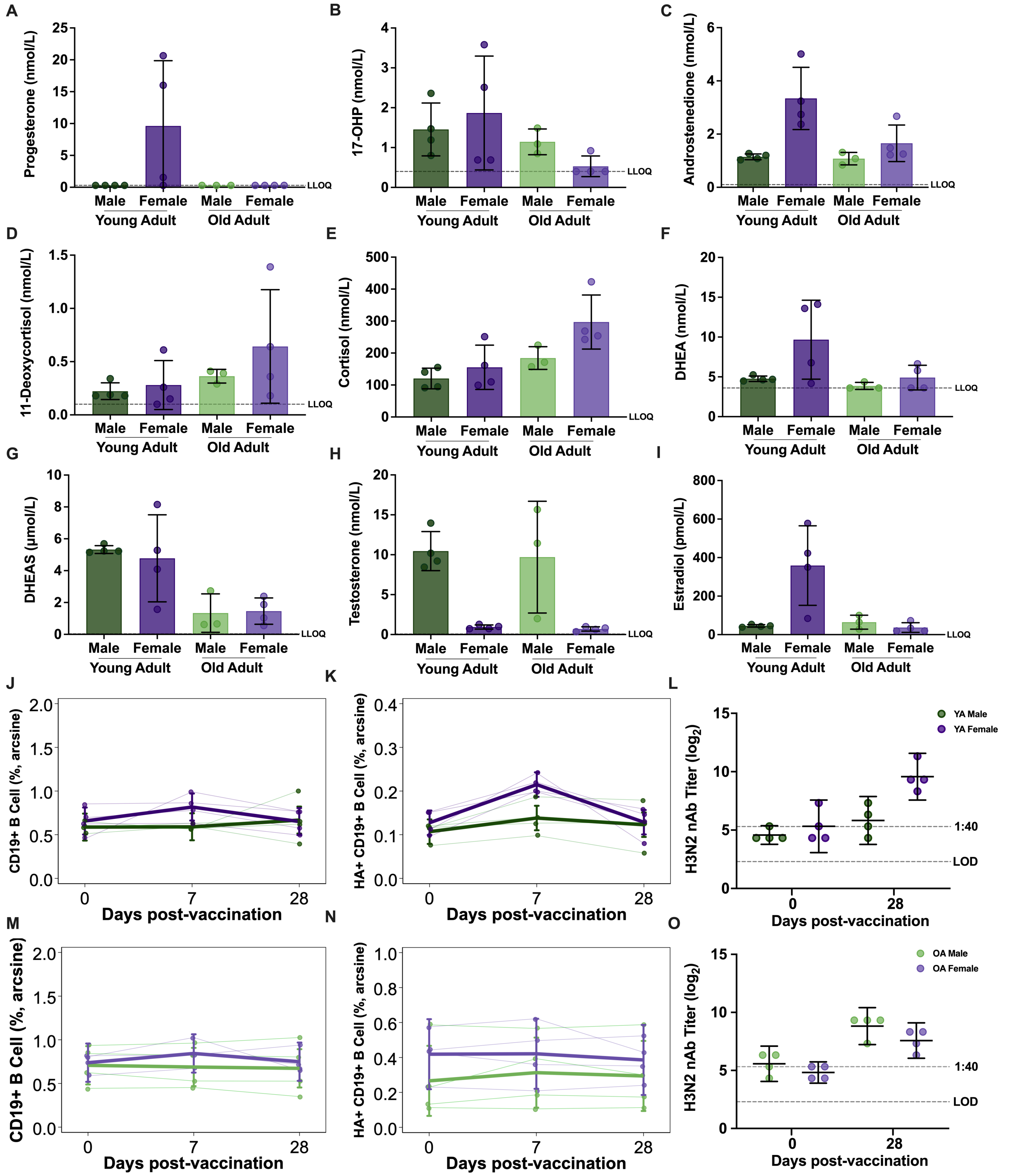

**Figure S8: Prior to vaccination, CD19+ B cells from young and old adults had distinct transcriptional profiles among male and female participants.** (**A**) Principal component analysis (PCA) of pseudobulk gene expression between young and old adults by sex. (**B**) Venn diagram of differentially expressed genes (DEGs) with padj<0.05 by sex between young and old adult participants. (**C-D**) Hallmark pathway gene set enrichment analysis (GSEA) of DEGs between young and old adult females or males, respectively, with bars labeled with the # of leading-edge genes for the top three significantly enriched pathways (padj<0.05). Top leading-edge genes from the TNF signaling (**E-F**), inflammatory response (**G-H**), and interferon alpha (**I-J**) enriched pathways that were shared between males and females in young (**E,G,I**) and old (**F,H,J**) adults are shown with padj<0.05 considered significant.

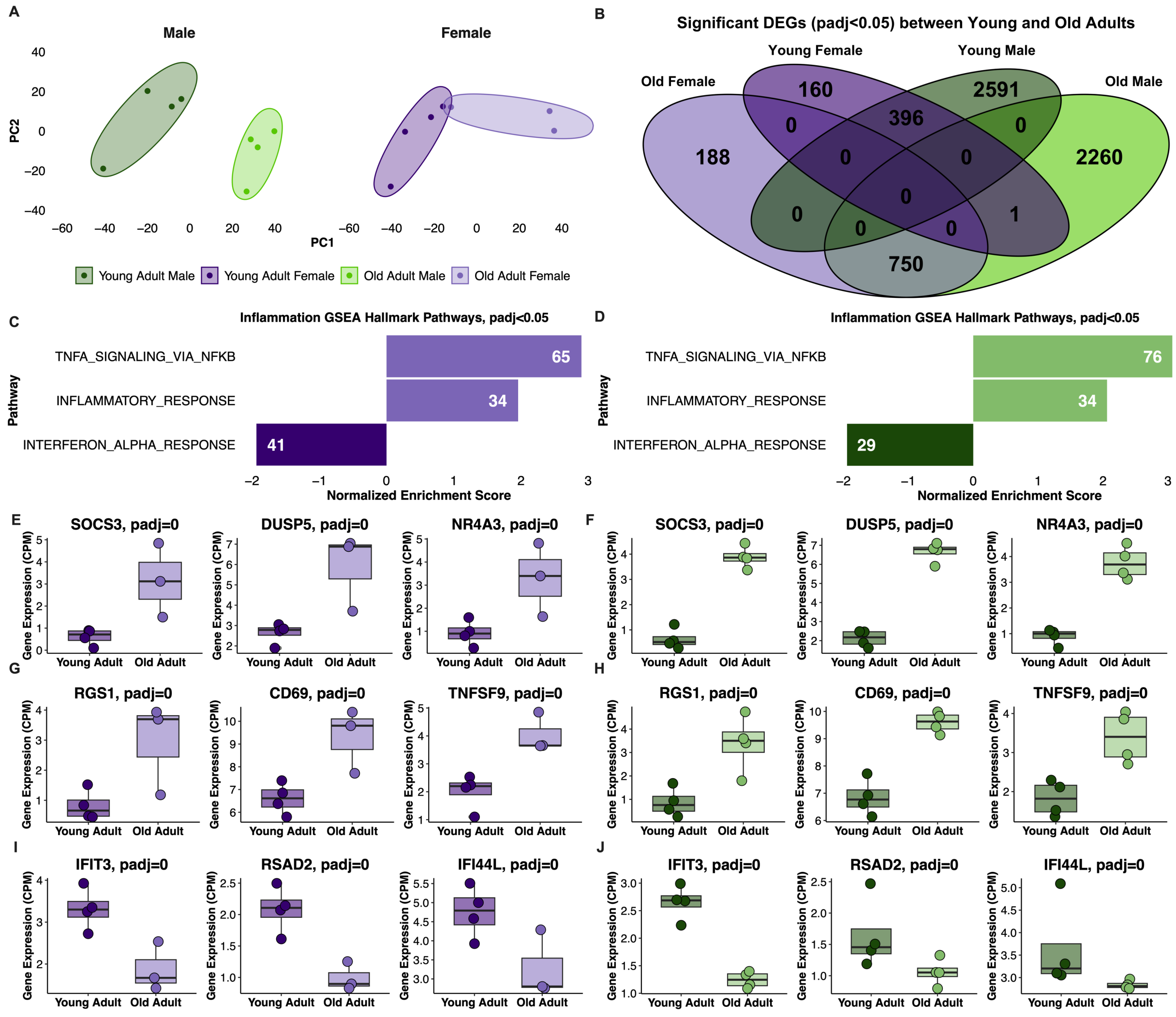

**Figure S9: Identification of B cell subsets from single cell RNA sequencing of FACS-sorted CD19+ B cells in peripheral mononuclear blood cells (PBMCs) collected from young and old adults. (A, C)** Clustering of single cells among young and old adults, respectively, using Seurat Uniform Manifold Approximation and Projection (UMAP) with manual cell type annotation of canonical markers (*10*). (**B, D**) Cell type markers used for manual cell type annotation for single cells of young and old adults, respectively. (**E-G**) Relative proportions of cell types for each sample used for single cell RNA-sequencing among young adults at 0-, 7-, and 28-days post-vaccination (DPV). (**H-J**) Relative proportions of cell types for each sample used for single cell RNA-sequencing among old adults at 0, 7, and 28 DPV.

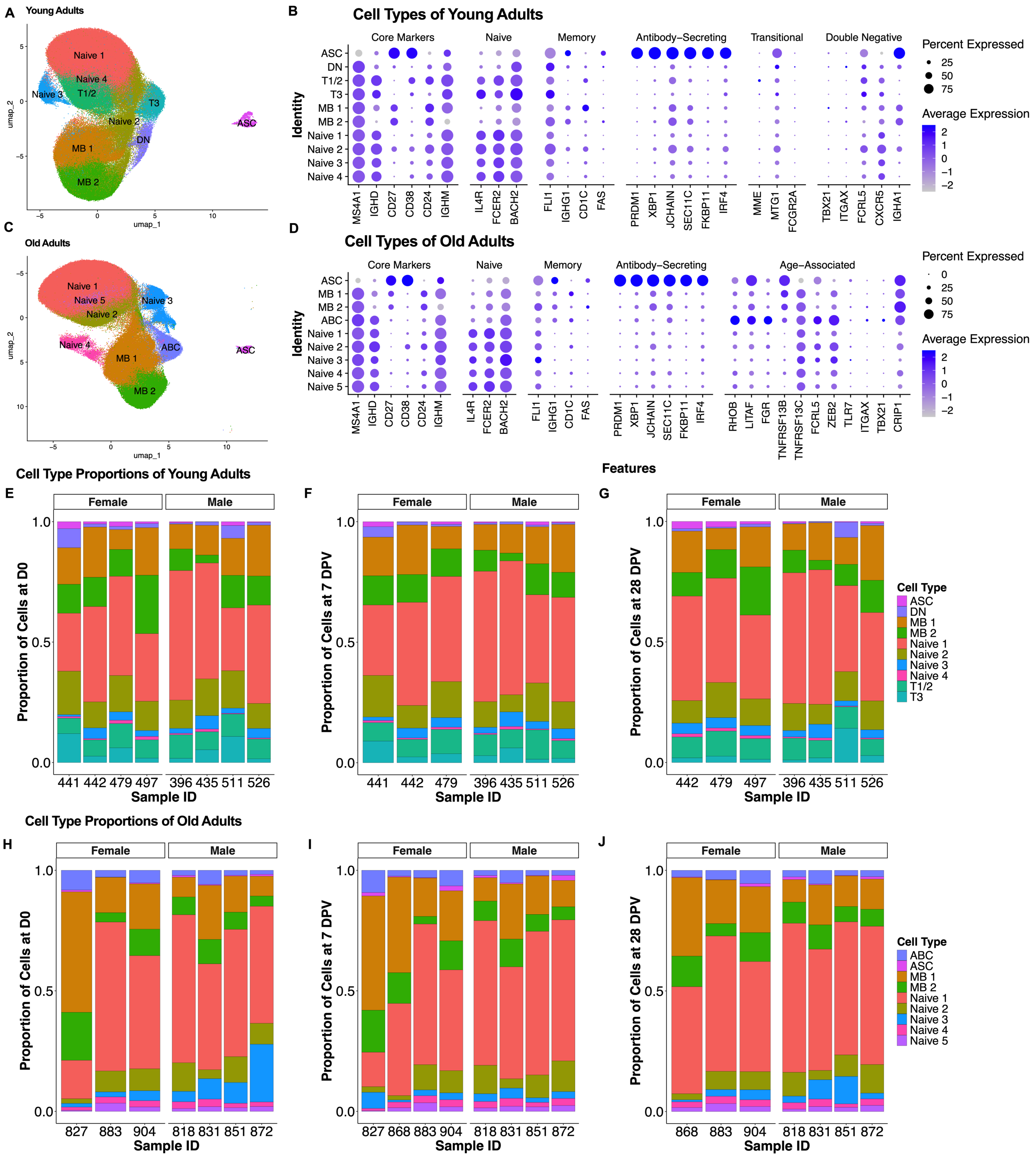

**Figure S10: Estrogen receptor signaling enhanced H3N2-, and to a lesser extent H1N1-specific IgG production in female antibody-secreting cells compared to males, independent of age.** PBMCs from equal numbers of male and female participants were analyzed for H1N1-specific IgG+ production (**A, C**) and H3N2-specific IgG+ production at multiple time points (**B, D**). Representative images of antibody-secreting cells (ASCs) producing HA-specific IgG+ from young adults (**A, B**) and old adults (**C, D**) from PBMCs collected from male and female participants prior to and 7- and 28-days post-vaccination (DPV). Spots represent H1N1- and H3N2-specific IgG+ cells. (**E–H**) Quantification of the number of H1N1- or H3N2-specific IgG+ cells in PBMCs from young adults (**E, F**) and old adults (**G, H**). (**I–L**) Quantification of spot sizes corresponding to H1N1- or H3N2-specific IgG+ in young (**I, J**) and old adults (**K, L**). A total of $1 \times 10^6$ PBMCs from 4 male and 4 female participants were used for all assays per group. Data were analyzed by two-way ANOVA. * represents $p < 0.05$.

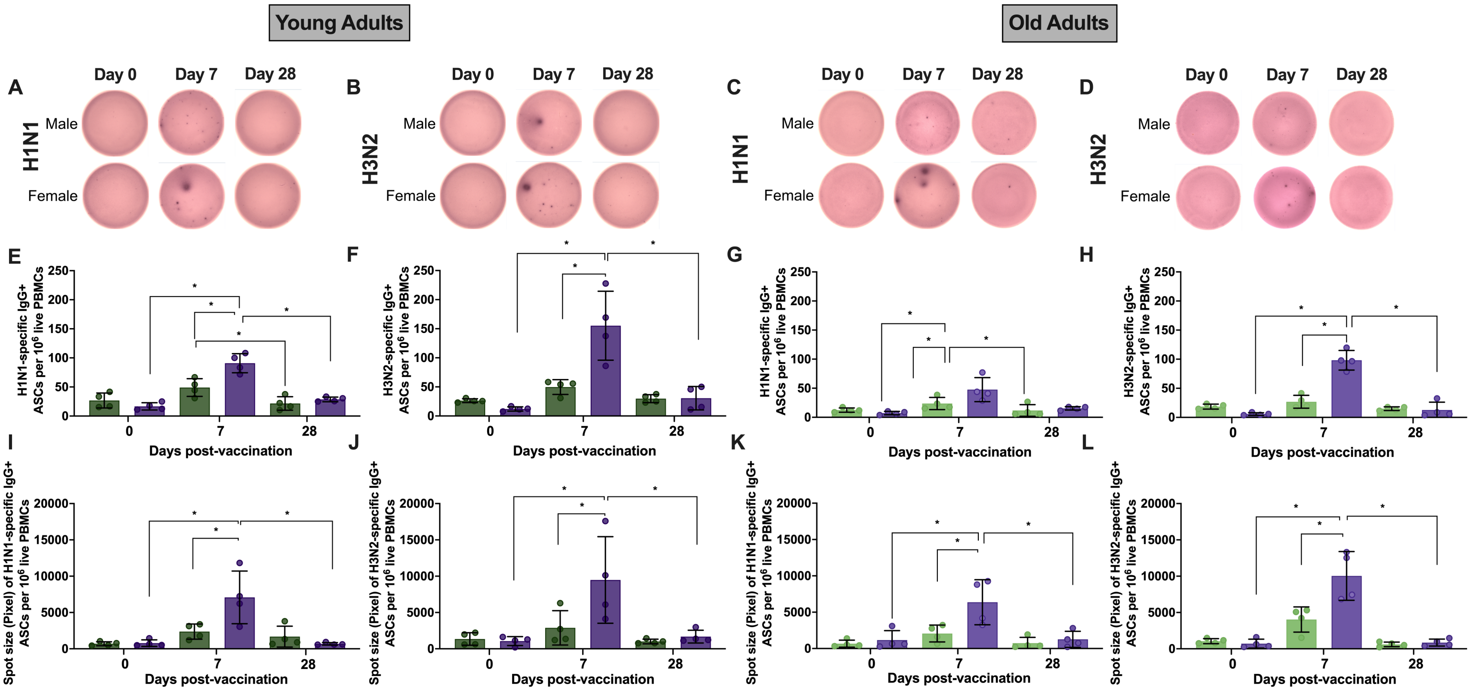

**Table S1: Summary of PBMC viability following ELISpot inhibitor treatments**

| | % cell viability | Average HA-specific IgG+ ASCs per $10^6$ live PBMCs | Average spot size (pixels) of HA-specific IgG+ ASCs per $10^6$ live PBMCs |
|---|---|---|---|
| Untreated | 92.5 | 102.94 | 7237.40 |
| E2 | 82.0 | 194.85 | 21713.77 |
| TAM | 80.1 | 78.02 | 7971.61 |
| NFkB inhibitor (1μM) | 80.2 | 79.04 | 3963.56 |
| NFkB inhibitor (1μM)+E2 | 81.5 | 64.34 | 3474.73 |
| NFkB inhibitor (5μM) | 84.0 | 51.47 | 1714.07 |
| NFkB inhibitor (5μM)+E2 | 85.9 | 51.47 | 1928.98 |
| NFkB inhibitor (10μM) | 60.7 | NA | NA |
| NFkB inhibitor (10μM)+E2 | 50.1 | NA | NA |
| TNF inhibitor (1μg/mL) | 86.0 | 55.15 | 2936.72 |
| TNF inhibitor (5μg/mL) | 83.2 | 31.25 | 2634.57 |
| TNF inhibitor (1μg/mL)+E2 | 85.3 | 49.63 | 1016.39 |
| TNF inhibitor (5μg/mL)+E2 | 82.3 | 35.85 | 1257.52 |

**Table S2: Flow cytometric primary antibodies used for characterizing human B cell subsets**

| Target Cell Population | Antibody | Fluorochrome | Clone | Dilution | Manufacturer | Cat. # |
|---|---|---|---|---|---|---|
| CD19+ B cell subpopulations (B cell, MB cell, ASC) | CD3 | BV786 | HIT3a | 1:100 | BD Biosciences | 740961 |
| | CD14 | BV786 | M5E2 | 1:100 | BD Biosciences | 563698 |
| | CD16 | BV786 | 3G8 | 1:100 | BD Biosciences | 563690 |
| | CD71 | APC | M-A712 | 1:100 | BD Biosciences | 551374 |
| | CD38 | BV605 | HIT2 | 1:100 | BD Biosciences | 740401 |
| | CD19 | BV421 | HIB19 | 1:100 | BD Biosciences | 562440 |
| | CD20 | PE-Cy7 | 2H7 | 1:100 | BD Biosciences | 560735 |
| | IgD | PerCP-Cy5.5 | IA6-2 | 1:100 | BD Biosciences | 561315 |
| ABC | CD3 | BV786 | HIT3a | 1:100 | BD Biosciences | 740961 |
| | CD14 | BV786 | M5E2 | 1:100 | BD Biosciences | 563698 |
| | CD16 | BV786 | 3G8 | 1:100 | BD Biosciences | 563690 |
| | CD19 | FITC | HIB19 | 1:100 | Fisher Scientific | BDB555412 |
| | CD21 | PE-Cy7 | B-ly4 | 1:100 | Fisher Scientific | BDB561374 |
| | T-bet | BV421 | O4-46 | 1:100 | Fisher Scientific | BDB563318 |
| | CD11c | BV605 | B-ly6 | 1:100 | Fisher Scientific | BDB563929 |
| HA+ cells | Steptavidin | PE | | 1:200 | BD Biosciences | 554061 |
| Live cells | Live/Dead | Aqua | | 1uL | Invitrogen | L34957 |
| | 7-AAD | 7-AAD | | 1ug/mL | Thermo Fisher | A1310 |

**Table S3: Cell markers of human total and HA-specific CD19+ B cell subsets used for flow cytometric analyses**

| Target Cell Population | Markers |
|---|---|
| CD19+ B cells | CD3e-CD14-CD16-CD19+ |
| HA+ CD19+ B cells | CD3e-CD14-CD16-CD19+H1+H3+ |
| ASCs | CD3e-CD14-CD16-CD19+IgD-CD71+CD38+CD20- |
| HA+ ASCs | CD3e-CD14-CD16-CD19+IgD-CD71+CD38+CD20-H1+H3+ |
| MB cells | CD3e-CD14-CD16-CD19+IgD-CD71- |
| HA+ MB cells | CD3e-CD14-CD16-CD19+IgD-CD71-H1+H3+ |
| ABCs | CD3e-CD14-CD16-CD19+CD21-CD11c+Tbet+ |
| HA+ ABCs | CD3e-CD14-CD16-CD19+CD21-CD11c+Tbet+H1+H3+ |

**Table S4: qPCR Primers**

| Gene name | Sequence |
|---|---|
| ACTB_F | 5'-GAAGATCAAGATCATTGCTCCT-3' |
| ACTB_R | 5'-TACTCCTGCTTGCTGATCCA-3' |
| TNFA_F | 5'-CCCAGGCAGTCAGATCATCTTC-3' |
| TNFA_R | 5'-AGCTGCCCCTCAGCTTGA-3' |
| SOCS3_F | 5'-TTTCTGATCCGCGACAGCT-3' |
| SOCS3_R | 5'-GGTCCCAGACTGGGTCTTGA-3' |
| SLC2A3_F | 5'-CACAACAGAGGCAAGGGTACATAT-3' |
| SLC2A3_R | 5'-AAGAAGGAGCAAATGCCAAGTT-3' |
| IRF4_F | 5'-GGATTGCTGATGTGTTCTGGTA-3' |
| IRF4_R | 5'-GGCAAAGAAAGCTCATCACAG-3' |

### SUPPLEMENTAL MATERIALS REFERENCES


1. H. Kuo, J. R. Shapiro, S. Dhakal, R. Morgan, A. L. Fink, H. Liu, J. W. Westerbeck, K. E. Sylvia, H. S. Park, R. L. Ursin, P. Shea, K. Shaw-Saliba, K. Fenstermacher, R. Rothman, A. Pekosz, S. L. Klein, Sex-specific effects of age and body mass index on antibody responses to seasonal influenza vaccines in healthcare workers. *Vaccine* **40**, 1634-1642 (2022).
2. T. Potluri, A. L. Fink, K. E. Sylvia, S. Dhakal, M. S. Vermillion, L. Vom Steeg, S. Deshpande, H. Narasimhan, S. L. Klein, Age-associated changes in the impact of sex steroids on influenza vaccine responses in males and females. *NPJ Vaccines* **4**, 29 (2019).
3. E. Faenzi, L. Zedda, M. Bardelli, F. Spensieri, E. Borgogni, G. Volpini, F. Buricchi, F. L. Pasini, P. L. Capecchi, F. Montanaro, R. Belli, M. Lattanzi, S. Piccirella, E. Montomoli, S. S. Ahmed, R. Rappuoli, G. Del Giudice, O. Finco, F. Castellino, G. Galli, One dose of an MF59-adjuvanted pandemic A/H1N1 vaccine recruits pre-existing immune memory and induces the rapid rise of neutralizing antibodies. *Vaccine* **30**, 4086-4094 (2012).
4. D. Frasca, A. Diaz, M. Romero, B. B. Blomberg, The generation of memory B cells is maintained, but the antibody response is not, in the elderly after repeated influenza immunizations. *Vaccine* **34**, 2834-2840 (2016).
5. Y. C. Lin, Y. C. Lin, C. C. Wu, M. Y. Huang, W. C. Tsai, C. H. Hung, P. L. Kuo, The immunomodulatory effects of TNF-alpha inhibitors on human Th17 cells via RORgammat histone acetylation. *Oncotarget* **8**, 7559-7571 (2017).
6. R. Miyamoto, T. Ito, S. Nomura, R. Amakawa, H. Amuro, Y. Katashiba, M. Ogata, N. Murakami, K. Shimamoto, C. Yamazaki, K. Hoshino, T. Kaisho, S. Fukuhara, Inhibitor of IkappaB kinase activity, BAY 11-7082, interferes with interferon regulatory factor 7 nuclear translocation and type I interferon production by plasmacytoid dendritic cells. *Arthritis Res Ther* **12**, R87 (2010).
7. S. J. E. Verdonk, H. W. Vesper, F. Martens, P. M. Sluss, J. J. Hillebrand, A. C. Heijboer, Estradiol reference intervals in women during the menstrual cycle, postmenopausal women and men using an LC-MS/MS method. *Clin Chim Acta* **495**, 198-204 (2019).
8. I. Korsunsky, N. Millard, J. Fan, K. Slowikowski, F. Zhang, K. Wei, Y. Baglaenko, M. Brenner, P. R. Loh, S. Raychaudhuri, Fast, sensitive and accurate integration of single-cell data with Harmony. *Nature methods* **16**, 1289-1296 (2019).
9. Y. Hao, T. Stuart, M. H. Kowalski, S. Choudhary, P. Hoffman, A. Hartman, A. Srivastava, G. Molla, S. Madad, C. Fernandez-Granda, R. Satija, Dictionary learning for integrative, multimodal and scalable single-cell analysis. *Nat Biotechnol* **42**, 293-304 (2024).
10. I. Sanz, C. Wei, S. A. Jenks, K. S. Cashman, C. Tipton, M. C. Woodruff, J. Hom, F. E. Lee, Challenges and Opportunities for Consistent Classification of Human B Cell and Plasma Cell Populations. *Front Immunol* **10**, 2458 (2019).
11. K. Blighe, S. M. Rana, M. Lewis. (2021).
12. A. Liberzon, C. Birger, H. Thorvaldsdottir, M. Ghandi, J. P. Mesirov, P. Tamayo, The Molecular Signatures Database (MSigDB) hallmark gene set collection. *Cell Syst* **1**, 417-425 (2015).
13. H. Chen, P. C. Boutros, VennDiagram: a package for the generation of highly-customizable Venn and Euler diagrams in R. *BMC Bioinformatics* **12**, 35 (2011).
14. N. Borcherding, A. Vishwakarma, A. P. Voigt, A. Bellizzi, J. Kaplan, K. Nepple, A. K. Salem, R. W. Jenkins, Y. Zakharia, W. Zhang, Mapping the immune environment in clear cell renal carcinoma by single-cell genomics. *Communications Biology* **4**, 122 (2021).
15. A. P. Anderson, A. G. Jones, erefinder: Genome-wide detection of oestrogen response elements. *Mol Ecol Resour* **19**, 1366-1373 (2019).
16. J. Peretz, A. Pekosz, A. P. Lane, S. L. Klein, Estrogenic compounds reduce influenza A virus replication in primary human nasal epithelial cells derived from female, but not male, donors. *Am J Physiol Lung Cell Mol Physiol* **310**, L415-425 (2016).
17. V. V. Tyulmenkov, C. M. Klinge, A mathematical approach to predict the affinity of estrogen receptors alpha and beta binding to DNA. *Mol Cell Endocrinol* **182**, 109-119 (2001).
18. Z. Li, T. Li, M. E. Yates, Y. Wu, A. Ferber, L. Chen, D. D. Brown, J. S. Carroll, M. J. Sikora, G. C. Tseng, S. Oesterreich, A. V. Lee, EstroGene database reveals diverse temporal, context-dependent and directional estrogen receptor regulomes in breast cancer. *bioRxiv*, (2023).

19. M. Kuhn, Building Predictive Models in R Using the caret Package. *Journal of Statistical Software* **28**, 1 - 26 (2008).
20. X. Robin, N. Turck, A. Hainard, N. Tiberti, F. Lisacek, J. C. Sanchez, M. Muller, pROC: an open-source package for R and S+ to analyze and compare ROC curves. *BMC Bioinformatics* **12**, 77 (2011).
